# CHANGES IN CERVICAL MUSCLE MECHANICAL PROPERTIES DURING OROFACIAL MOTOR TASKS IN INDIVIDUALS WITH AND WITHOUT CHRONIC NECK PAIN

**DOI:** 10.64898/2026.11.12.26367942

**Authors:** Alberto Pérez-González, Javier Sánchez-Palacios, Paula Fernández-González, Alba París-Alemany, Jorge Chamorro-Sánchez, Roy La Touche

## Abstract

**Objectives:** To investigate changes in cervical muscle mechanical properties during tongue-pressure tasks and maximum intercuspation (MIC) in individuals with chronic neck pain and healthy controls, and to explore associations between these properties, cervical mobility, tongue pressure, pain intensity, and neck disability.

**Methods:** Eighty-four adults participated, including 42 individuals with chronic neck pain and 42 healthy controls. Myotonometry assessed frequency, stiffness, logarithmic decrement, relaxation time, and creep in the sternocleidomastoid (SCM) and trapezius muscles during tongue-pressure tasks at 50%, 80%, and 100% of maximum tongue pressure (MTP) and during maximum intercuspation. Group-by-condition effects were examined using linear mixed-effects models. Exploratory correlations and cervical mobility assessments were conducted in a subsample of 36 participants.

**Results:** Tongue-pressure conditions showed significant effects on cervical muscle mechanical properties in 19 of 20 mixed-effects models at the nominal significance level, particularly for frequency, stiffness, relaxation time, and creep. Maximum intercuspation was associated with changes in frequency and stiffness in both muscles, with additional changes in relaxation time and creep in the sternocleidomastoid. Group-by-condition interactions were generally nonsignificant, apart from an isolated interaction involving right trapezius logarithmic decrement. The neck pain group exhibited lower absolute tongue-pressure values at 80% and 100% of maximum tongue pressure. Exploratory analyses identified reduced cervical extension and rotation, together with associations between cervical mobility, myotonometric properties, pain intensity, and tongue pressure.

**Conclusions:** Orofacial motor tasks were associated with changes in cervical muscle mechanical properties regardless of chronic neck pain status. Although clinical characteristics were associated with selected mechanical and functional measures, chronic neck pain did not consistently modify task-related cervical responses. These findings support functional interactions between cervical and orofacial systems.

## 1. INTRODUCTION

The relationship between cervical and orofacial function has been investigated through clinical observations, experimental studies of motor coordination, and assessments of the anatomical and neurophysiological connections between these regions. A systematic review and meta-analysis by Bizzarri et al. (2026) found an association between neck pain and temporomandibular disorders, although the available evidence was limited by methodological heterogeneity and low certainty. Research examining craniocervical posture has also reported relationships with temporomandibular symptoms and mandibular function (Armijo-Olivo et al., 2011; Ozmen & Unuvar, 2025). These associations are clinically relevant, but their interpretation requires caution because the coexistence of cervical and temporomandibular symptoms does not establish a common mechanical origin or indicate that dysfunction in one region necessarily produces dysfunction in the other.

Experimental investigations provide a more specific account of how cervical and mandibular motor behavior may be coordinated. Yamada et al. (1999) examined the influence of head posture on the direction and stability of mandibular closing movements. Eriksson et al. (2000) and Zafar et al. (2000) subsequently described temporal relationships between mandibular and head-neck movements during rhythmic jaw activities. These findings indicate that mandibular movement is accompanied by coordinated adjustments involving the cervical region, although movement coordination cannot be assumed to reflect a corresponding change in the mechanical properties of individual cervical muscles. Clinical investigations have also examined the influence of craniocervical posture on mandibular opening and pressure pain sensitivity in patients with myofascial temporomandibular disorders (La Touche et al., 2011), as well as the effects of cervical interventions on pain-related outcomes in this population (La Touche et al., 2009). The findings support the clinical relevance of cervical-orofacial interactions without establishing the mechanisms responsible for the observed associations.

Several anatomical and neurophysiological relationships may contribute to coordination between these regions. The tongue, mandible, and hyoid system form an interconnected functional arrangement in which lingual movements occur alongside adjustments of adjacent structures (Messina, 2017). Coordination between masticatory and tongue muscle activity has been examined during chewing tasks involving different food consistencies (Kakizaki et al., 2002). Neurophysiologically, the trigeminocervical complex provides a basis for convergence between trigeminal and upper cervical sensory pathways (Bartsch & Goadsby, 2003), and functional neuroimaging has contributed evidence concerning its representation in the human brainstem (Mehnert et al., 2023). These findings are relevant to the processing of craniofacial and cervical sensory information, but they do not directly demonstrate the motor mechanisms underlying cervical muscle responses during tongue-pressure tasks. Swallowing, in particular, depends on coordinated neural control involving multiple oropharyngeal structures (Costa, 2018), making it necessary to distinguish the physiological organization of swallowing from the more restricted motor demands imposed by experimental tongue-pressure tasks.

The relationship between cervical positioning and lingual function has also been examined through studies of swallowing biomechanics and tongue-pressure performance. Investigations involving cervical spine disorders have described associations with dysphagia and alterations in swallowing function (Abdel-Aziz et al., 2018; Papadopoulou et al., 2013; Walters et al., 2023). Experimental and clinical studies have additionally evaluated the influence of head and neck positioning on oropharyngeal swallowing (Ertekin et al., 2001; Perry et al., 2012; Solazzo et al., 2012). Electromyographic investigations of the suprahyoid and infrahyoid musculature have examined relationships between swallowing-related muscle activity and craniofacial morphology (Trevisan et al., 2013). Although these studies establish a basis for considering cervical positioning in the assessment of oropharyngeal function, their findings cannot be directly extrapolated to maximal voluntary tongue-pressure generation, which differs from swallowing in its motor requirements and temporal organization.

Studies using the Iowa Oral Performance Instrument have examined the effects of experimentally modified craniocervical conditions on tongue strength and endurance. Paris-Alemany et al. (2021) assessed tongue performance in neutral, forward, and retracted head positions and identified differences in tongue strength associated with craniocervical posture, particularly in the anterior and middle regions of the tongue. Tongue endurance also varied according to the region assessed, although differences between cervical positions were less consistent. Pérez-González et al. (2025) subsequently investigated tongue mobility and strength during passive cervical immobilization. Their findings suggested that restricting cervical movement may influence selected aspects of lingual performance, with a potentially greater effect in the middle region of the tongue. However, the absence of a consistent overall effect on tongue strength indicates that the relationship cannot be reduced to a uniform loss of force under cervical restriction. Differences between these investigations may reflect the distinct mechanical constraints imposed by changes in head position and passive immobilization, as well as the regional characteristics of the tongue measurements.

These experiments have primarily examined how cervical conditions influence lingual performance. The complementary question concerns the mechanical behavior of cervical muscles while lingual force is being generated. This distinction matters because a change in tongue-pressure output under altered cervical conditions does not establish whether cervical muscles undergo measurable mechanical adjustments during the task itself. Such responses could depend on muscle recruitment, stabilization demands, or changes in the mechanical loading of superficial tissues. Distinguishing among these explanations requires measurements beyond tongue-pressure performance alone.

Functional relationships involving the cervical region are not limited to lingual tasks. Electromyographic studies have reported associations between sternocleidomastoid (SCM) and masseter muscle activity under different occlusal conditions (Miralles et al., 1992), whereas analyses of mandibular kinematics have revealed coordinated jaw and head-neck movements (Eriksson et al., 2000; Zafar et al., 2000). These observations support the existence of functional coupling between the masticatory and cervical systems and justify the investigation of cervical muscle behaviour maximum intercuspation. Nevertheless, electromyographic activity and muscle mechanical properties reflect different aspects of neuromuscular function. Consequently, variations in electrical activation do not necessarily reflect corresponding changes in stiffness, oscillation frequency, or viscoelastic behavior, highlighting the need for direct assessment of the mechanical response to maximal dental clenching.

Myotonometry offers a means of characterizing the mechanical behavior of superficial muscles through parameters including oscillation frequency, stiffness, logarithmic decrement, relaxation time, and creep. Magdič et al. (2025) investigated acute myotonometric changes in the masseter and upper trapezius muscles following exercise and chewing tasks, reporting parameter-dependent responses that differed between the muscles and experimental activities. Their measurements were obtained before and after the interventions, leaving the mechanical behavior of cervical muscles during sustained orofacial contractions less clearly characterized. Della Posta et al. (2025) examined biomechanical and viscoelastic properties of masticatory muscles in individuals with temporomandibular disorders using MyotonPRO measurements. This work supports the application of myotonometry to the assessment of orofacial musculature, although the clinical interpretation of individual parameters remains dependent on their measurement properties and the conditions under which recordings are obtained. Myotonometric measurements describe tissue mechanical behavior and cannot independently determine the neural drive or electrical activation responsible for changes observed during a motor task.

The presence of chronic neck pain introduces a further question concerning the potential modification of these mechanical responses. Pain-related adaptations in cervical motor behavior could influence how the SCM and upper trapezius muscles respond to orofacial demands, but the direction and magnitude of such differences cannot be assumed from the coexistence of cervical and temporomandibular symptoms. The associations reported in clinical populations, including those described by Bizzarri et al. (2026), concern different outcomes from the task-related mechanical properties examined in the present study. Whether individuals with chronic neck pain exhibit distinct cervical myotonometric responses during tongue-pressure generation or maximum intercuspation (MIC) therefore remains an empirical question.

Relationships between cervical and orofacial function may also extend beyond responses to individual motor tasks. Investigations of swallowing-related musculature have reported associations with trunk muscle characteristics (Yamaguchi et al., 2023), while Amornsuradech et al. (2024) examined age-related differences in perioral and trunk muscle characteristics. Nakamura et al. (2024) investigated the relationship between seated spinal sagittal alignment and swallowing speed in older women. These studies concern populations and functional outcomes that differ from those examined here, but they illustrate the relevance of considering musculoskeletal characteristics outside the immediate orofacial region. In the present study, the relationships of interest involve cervical mobility, tongue-pressure performance, cervical muscle mechanical properties, and clinical characteristics. Their examination may help characterize associations within the cervical-orofacial system, although cross-sectional correlations cannot establish the direction of these relationships.

The present study had three objectives. The first was to investigate changes in the mechanical properties of the SCM and upper trapezius muscles during tongue-pressure tasks performed at 50%, 80%, and 100% of maximum tongue pressure (MTP), including whether these responses differed between individuals with chronic neck pain and healthy controls. The second objective was to examine cervical muscle mechanical responses during MIC and compare these responses between groups. The third was to explore associations between cervical mobility and function, tongue-pressure performance, cervical muscle mechanical properties, and clinical characteristics.

We hypothesized that tongue-pressure tasks would be accompanied by changes in cervical muscle mechanical properties, with responses potentially varying according to pressure intensity and clinical group. MIC was also expected to be associated with changes in cervical mechanical properties, although the available evidence did not support a specific directional prediction for between-group differences. For the exploratory analyses, we anticipated associations between selected measures of cervical mobility, tongue-pressure performance, and myotonometric properties, with possible relationships involving pain-related clinical characteristics in the neck pain group.

## 2. MATERIALS AND METHODS

### Study design

A non-randomized laboratory study with a mixed repeated-measures design was conducted to investigate cervical muscle mechanical responses during standardized orofacial motor tasks in individuals with chronic neck pain and healthy controls, following the Strengthening the Reporting of Observational Studies in Epidemiology (STROBE) guidelines (*von Elm et al., 2014*). Participants were classified into two pre-existing clinical groups, while experimental conditions were assessed repeatedly within participants.

The study addressed three objectives. The first was to examine changes in the mechanical properties of the upper trapezius and SCM muscles during tongue-pressure tasks performed at 50%, 80%, and 100% of individual MTP, and to determine whether these responses differed between groups. The second was to investigate cervical muscle mechanical responses during MIC and compare these responses between groups. The third was to explore associations among cervical range of motion, MTP, cervical muscle mechanical properties, and clinical characteristics.

Cervical muscle mechanical properties were assessed using myotonometry, including oscillation frequency, dynamic stiffness, logarithmic decrement, relaxation time, and creep. Cervical mobility was evaluated using a Cervical Range of Motion (CROM) device. Pain intensity and neck-related disability were assessed in participants with chronic neck pain.

The association analyses were exploratory and were conducted using the available measurements from the complementary assessment. These analyses were intended to characterize relationships between variables rather than establish causal mechanisms.

The study protocol was approved by the Research Ethics Committee of the Centro Superior de Estudios Universitarios La Salle (CSEULS-PI 010/2020). All procedures were conducted in accordance with the Declaration of Helsinki.

### Participants

A total of 84 participants, aged 18–65 years, were included in the study. Participants were recruited through convenience sampling from specialized clinical settings and the university campus of the Centro Superior de Estudios Universitarios La Salle. Based on their clinical characteristics and predefined eligibility criteria, participants were classified into two groups: a control group (CG; n = 42) and a neck pain group (NPG; n = 42).

The CG comprised asymptomatic individuals who had not experienced pain or dysfunction involving the craniocervical, temporomandibular, or swallowing regions during the preceding 12 months. Individuals with a history of recent trauma or surgery affecting these regions were not eligible for inclusion.

The NPG comprised individuals presenting with mechanical neck pain, defined by pain intensity greater than 3 points on the Numeric Rating Scale (NRS) and neck-related disability greater than 5 points on the Neck Disability Index (NDI). Group classification was based on clinical screening and the application of these eligibility criteria.

General exclusion criteria for both groups included diagnosed neuromuscular, neurological, or systemic disorders potentially affecting pain perception; trauma or surgery involving the craniocervical region within the preceding year; recent use of analgesic or anti-inflammatory medication; pregnancy or lactation; and concurrent participation in other research studies.

Eligibility screening and participant assessment were conducted by a physiotherapist with six years of clinical experience in the management of craniofacial disorders and chronic pain.

### Variables and instruments

#### - Numeric Rating Scale (NRS)

Pain intensity was assessed in the neck pain group using the Numeric Rating Scale (NRS), an 11-point numerical scale ranging from 0 (no pain) to 10 (worst imaginable pain). Participants were asked to select the number that best represented their neck pain intensity during the preceding week. The NRS was used to characterize pain severity and determine eligibility for the neck pain group, for which a score greater than 3 was required. Numerical pain rating scales are widely used in musculoskeletal pain research and have demonstrated acceptable measurement properties, although these properties may vary according to the population and assessment context (Chiarotto et al., 2019).

#### - Neck Disability Index (NDI)

Neck-related disability was assessed using the Spanish version of the Neck Disability Index (NDI) (Andrade Ortega et al., 2010). The questionnaire comprises 10 items addressing the impact of neck pain on daily activities and functioning. Each item is scored from 0 to 5, yielding a total score ranging from 0 to 50, with higher scores indicating greater disability. Scores can also be expressed as percentages by multiplying the total score by two. An NDI score greater than 5 was required for inclusion in the neck pain group. The NDI is commonly used to assess neck-related disability, and its measurement properties have been examined in systematic reviews of disability questionnaires for individuals with neck pain (Lemeunier et al., 2019).

#### - Cervical Range of Motion

Active cervical range of motion was assessed using the Cervical Range of Motion (CROM) device (Performance Attainment Associates, Roseville, MN, USA), a head-mounted instrument designed to quantify cervical mobility in degrees. The device consists of a lightweight frame resembling eyeglasses, supported over the bridge of the nose and ears and secured around the head with an adjustable strap. It incorporates three angular measurement systems that allow cervical movements to be assessed in the sagittal, frontal, and transverse planes.

Two gravity-dependent inclinometers measure cervical flexion and extension in the sagittal plane and lateral flexion in the frontal plane. Cervical rotation is measured using a magnetic inclinometer positioned on the upper part of the frame, which operates in conjunction with a magnetic reference yoke placed over the shoulders. This arrangement provides a reference for measuring head rotation relative to the upper trunk.

Six active cervical movements were assessed: flexion, extension, right and left lateral flexion, and right and left rotation. Each movement was expressed in degrees and considered a separate measure of cervical mobility.

The CROM device has demonstrated satisfactory measurement properties in previous investigations. Audette et al. (2010) reported good to excellent between-day test-retest reliability across the six cervical movements, with intraclass correlation coefficients (ICCs) ranging from 0.89 to 0.98. The standard error of measurement ranged from 1.6° to 2.8°, while the minimal detectable change ranged from 3.6° to 6.5°. In individuals with cervical dysfunction, Rheault et al. (1992) reported inter-rater reliability coefficients ranging from 0.76 to 0.98.

#### - Cervical Muscle Mechanical Properties

The mechanical properties of the cervical muscles were assessed using the MyotonPRO device (Myoton AS, Estonia), a non-invasive instrument that applies a brief mechanical impulse to the tissue through a probe positioned perpendicular to the skin surface. The resulting damped oscillation is recorded and used to calculate parameters describing the mechanical response of the underlying tissue.

Five myotonometric parameters were evaluated:

- Oscillation frequency (Hz): The natural frequency of the tissue oscillation induced by the mechanical impulse. This parameter reflects the oscillatory response of the tissue under the measurement conditions and is commonly used as an indirect indicator of mechanical tension.
- Dynamic stiffness (N/m): The resistance of the tissue to deformation induced by the mechanical impulse. Higher values indicate greater resistance to the applied mechanical displacement.
- Logarithmic decrement (dimensionless): A parameter describing the damping of the induced tissue oscillation. Higher values indicate greater oscillation damping and are conventionally interpreted as reflecting lower tissue elasticity.
- Mechanical stress relaxation time (ms): The time-related parameter describing the reduction in mechanical stress following deformation. It characterizes one aspect of the tissue’s viscoelastic response.
- Creep (dimensionless): A device-derived viscoelastic parameter representing the relationship between relaxation time and deformation time. It provides an additional description of time-dependent tissue mechanical behavior.

These parameters were considered measures of tissue mechanical behavior rather than direct indicators of muscle electrical activity or neural activation. Myotonometry has been applied to the assessment of masticatory and cervical musculature under different clinical and experimental conditions (Della Posta et al., 2025; Magdič et al., 2025).

Measurements were obtained bilaterally from the upper trapezius and SCM muscles both at rest and during active orofacial motor tasks.

#### - Tongue Strength and Endurance

Tongue pressure was assessed using the Iowa Oral Performance Instrument (IOPI; IOPI Medical LLC, USA), which measures the pressure generated by the tongue against an air-filled bulb positioned against the palate. Pressure values were expressed in kilopascals (kPa).

MTP was determined during a voluntary tongue-pressing task and used as the reference value for the experimental conditions. Participants subsequently performed tongue-pressure tasks corresponding to 100%, 80%, and 50% of their individual MTP. For the submaximal conditions, the IOPI provided visual feedback to assist participants in reaching and maintaining the target pressure during myotonometric measurements.

Tongue-pressure performance was treated as a measure of pressure-generating capacity. The experimental protocol did not include a separate standardized test of tongue endurance. The positioning of the pressure bulb, task instructions, and sequence of experimental conditions are described in the following section.

### Experimental procedures

#### Participant assessment and preparation

Participants who met the eligibility criteria received verbal and written information about the study objectives and experimental procedures. Written informed consent was obtained before any study-related assessments were performed.

Sociodemographic information was collected from all participants. In the NPG, pain intensity and neck-related disability were assessed using the NRS and NDI, respectively.

The experimental protocol was designed to assess the mechanical properties of the upper trapezius and SCM muscles at rest and during standardized orofacial motor tasks. Following the initial assessment, participants underwent resting myotonometry and four active motor conditions administered in a fixed sequence.

#### Experimental positioning and anatomical landmarks

All measurements were performed in a designated examination room under standardized conditions. Participants were seated with their backs supported, feet resting on the floor, legs uncrossed and positioned approximately hip-width apart, and the cervical spine maintained in a neutral position.

Before myotonometric assessment, the anatomical measurement sites were identified and marked on the skin using a dermographic marker. These markings were maintained throughout the experimental procedure to ensure consistent probe placement.

The two cervical muscles assessed at rest and during the active motor conditions were:

- Upper trapezius: The measurement site was located at the midpoint of the line connecting the spinous process of the seventh cervical vertebra (C7) and the lateral border of the acromion. Measurements were obtained bilaterally.
- Sternocleidomastoid: The measurement site was identified at the midpoint of the muscle belly along the line extending from the mastoid process toward the jugular notch of the sternum. Measurements were obtained bilaterally.

The side assessed first was determined by coin toss. This procedure randomized the initial measurement side but did not alter the fixed sequence of motor conditions.

#### Myotonometric measurements and quality control

Myotonometric measurements were obtained using the MyotonPRO in multiscan mode, with three consecutive mechanical impulses delivered during each measurement series. The probe was positioned perpendicular to the skin at the previously marked anatomical site.

A coefficient of variation below 5% was required for a measurement series to be accepted. Series exceeding this threshold were repeated. Two valid series were obtained for each measurement site and experimental condition, and the mean of the two series was used for statistical analysis.

At rest, during maximum intercuspation, and under all tongue-pressure conditions, myotonometric measurements were obtained bilaterally from the right and left upper trapezius and SCM muscles. Active measurements of the masticatory and suprahyoid muscles were not included in the experimental design because preliminary recordings during contraction showed coefficients of variation exceeding the predefined 5% acceptance threshold.

#### Maximum intercuspation

After the resting measurements, participants performed MIC as the first active motor condition. They were instructed to clench their teeth as firmly as possible while maintaining the standardized seated position.

During this task, myotonometric measurements were obtained from four cervical measurement sites: the right upper trapezius, left upper trapezius, right SCM, and left SCM. Two valid measurement series were obtained at each site.

#### Tongue-pressure tasks

Following maximum intercuspation, participants performed tongue-pressure tasks using the Iowa Oral Performance Instrument (IOPI). Maximum voluntary tongue pressure was established as the reference value for the subsequent submaximal conditions.

The tongue-pressure tasks were administered in the following fixed order: 100%, 80%, and 50% of individual MTP. During the maximal condition, participants pressed the IOPI bulb against the palate with maximum voluntary effort. During the 80% and 50% conditions, visual feedback from the device was used to help participants reach and maintain the corresponding target pressure.

At each pressure level, myotonometric measurements were obtained bilaterally from the upper trapezius and SCM muscles. Thus, the same four cervical measurement sites were assessed during MIC and each of the three tongue-pressure conditions.

Brief rest periods were provided between motor conditions to reduce the potential influence of accumulated fatigue. Participants maintained the standardized seated position throughout the experimental protocol.

#### Cervical range of motion assessment

Active cervical range of motion was assessed using the Cervical Range of Motion (CROM) device (Performance Attainment Associates, Roseville, MN, USA). Participants were seated with their backs supported, feet resting on the floor, and the cervical spine initially maintained in a neutral position.

The device was positioned on the participant’s head, with the anterior support resting on the bridge of the nose and the lateral supports positioned over the ears. An adjustable strap was used to secure the instrument and minimize displacement during cervical movements. For the assessment of cervical rotation, the magnetic reference yoke was positioned over the shoulders.

Before each measurement, the examiner verified the alignment of the instrument and established the neutral reference position. Participants were instructed to perform active cervical movements while maintaining their trunk position to minimize compensatory movements.

Six cervical movements were assessed: flexion, extension, right lateral flexion, left lateral flexion, right rotation, and left rotation. Flexion, extension, and lateral flexion were measured using the gravity-dependent inclinometers, whereas cervical rotation was assessed using the magnetic inclinometer in conjunction with the reference yoke.

Two consecutive measurements were obtained for each of the six cervical movements. Angular displacement was recorded in degrees, and the mean of the two measurements was calculated for each movement and used in the statistical analyses.

The resulting cervical mobility variables were included in the exploratory analyses examining associations among cervical range of motion, MTP, the mechanical properties of the upper trapezius and SCM muscles, and clinical characteristics.

#### Sample size calculation

Sample size estimation was performed using G*Power software (version 3.1.9.7; Heinrich Heine University Düsseldorf, Germany). The calculation was based on a repeated-measures analysis of variance (ANOVA), considering the interaction between group (individuals with chronic neck pain and healthy controls) and experimental condition. Given the limited evidence regarding cervical muscle mechanical responses during tongue-pressure tasks, a medium effect size (f = 0.25) was assumed. The significance level was set at α = 0.05, with a statistical power of 80% (1 − β = 0.80). The final study sample comprised 84 participants, with 42 individuals in each group.

In addition to the experimental assessment of cervical muscle mechanical properties during orofacial motor tasks, a complementary assessment was conducted to examine cervical range of motion using the Cervical Range of Motion (CROM) device and to explore associations among cervical mobility, MTP, muscle mechanical properties, and clinical characteristics.

These association analyses were considered exploratory. The sample size calculation was based on the primary repeated-measures design rather than on the detection of specific correlation coefficients. Accordingly, the exploratory findings were interpreted without assuming that the study was specifically powered to detect associations of a predetermined magnitude.

### Statistical Analysis

#### Descriptive analyses and preliminary data screening

Demographic and clinical characteristics were summarized using means and standard deviations for continuous variables and absolute frequencies and percentages for categorical variables. Between-group differences in continuous characteristics were assessed using Welch’s independent-samples t-test, with degrees of freedom estimated using the Welch–Satterthwaite approximation. Mean differences and 95% confidence intervals (CIs) were calculated for the CG and the NPG. Standardized between-group differences were expressed as Hedges’ g, incorporating a small-sample bias correction. Categorical variables were compared using Pearson’s chi-square test or Fisher’s exact test, as appropriate. Fisher’s exact test was applied to employment and marital status because of the distribution of observations across categories. Cramér’s V was calculated to quantify the association between group membership and sex. Pain intensity and neck disability were summarized exclusively for participants with chronic neck pain. Baseline comparisons were interpreted descriptively rather than as evidence of group equivalence.

Before inferential analyses, myotonometric measurements were examined according to group, experimental condition, muscle, anatomical side, and mechanical property. Distributional characteristics were evaluated using the mean, standard deviation, median, interquartile range, skewness, kurtosis, and visual inspection of quantile–quantile plots. Potential outliers were identified using the 1.5 × interquartile range criterion and examined against the available measurement records to identify possible transcription errors. Verified transcription errors were corrected before analysis, whereas valid extreme observations were retained. Differences in variance between groups were examined using the Brown–Forsythe test, based on deviations from group medians. The corresponding p-values were additionally evaluated using the Benjamini–Hochberg false discovery rate procedure. These preliminary assessments were used to characterize the data and inform the evaluation of model assumptions rather than to determine whether observations should be excluded.

#### Myotonometric responses to tongue-pressure tasks

Linear mixed-effects models were fitted to examine the effects of group and tongue-pressure condition on five myotonometric properties: muscle frequency, stiffness, logarithmic decrement, relaxation time, and creep. Separate models were estimated for the right and left SCM and trapezius muscles, yielding 20 models. Group (CG and NPG), experimental condition (rest and 50%, 80%, and 100% of MTP), and their interaction were specified as fixed effects. Experimental condition was treated as a categorical factor, and a participant-specific random intercept was included to account for repeated measurements within individuals.

Alternative residual variance structures were examined to accommodate potential heterogeneity across group-by-condition combinations. Models assuming homogeneous residual variances were compared with models allowing heterogeneous residual variances using maximum likelihood estimation and the Akaike information criterion (AIC). The heterogeneous structure was selected when its AIC was at least two units lower than that of the homogeneous model; otherwise, the homogeneous structure was retained. This criterion was used as a prespecified model-selection rule rather than as a formal significance test. The selected model was subsequently refitted using restricted maximum likelihood estimation. Model adequacy was evaluated through visual inspection of residual-versus-fitted plots, residual dispersion across model strata, and quantile–quantile plots of residuals. Age and sex were not included as covariates in the final models.

Fixed effects were evaluated using F-tests, and their magnitudes were summarized using partial eta squared (η²p), calculated from the F-statistic and its numerator and denominator degrees of freedom as η²p = (F × df₁)/(F × df₁ + df₂). Omnibus p-values were reported without adjustment across the 20 separately fitted models.

Post hoc comparisons were performed using estimated marginal means derived from the fitted mixed-effects models. Within-group contrasts compared each tongue-pressure condition with rest. Between-group contrasts examined differences between CG and NPG within each experimental condition. Additional difference-in-change contrasts compared condition-related changes from rest between groups. Holm’s step-down procedure was applied separately to the three within-group comparisons against rest for each group and muscle-property combination, to the four between-group comparisons across experimental conditions for each muscle-property combination, and to the three between-group difference-in-change contrasts for each muscle-property combination. Statistical significance was determined using Holm-adjusted p-values. The corresponding 95% confidence intervals were calculated without multiplicity adjustment and were therefore interpreted separately from the adjusted p-values.

The standardized magnitude of within-group changes relative to rest was described using Hedges’ g_av, based on the observed paired measurements and standardized using the square root of the average of the variances at rest and during the corresponding tongue-pressure condition. A small-sample bias correction was applied. These descriptive standardized effect sizes were distinguished from the inferential estimates obtained from the mixed-effects models. Accordingly, the statistical significance of within-group changes was determined from model-based contrasts rather than from the standardized effect sizes.

#### Myotonometric responses to maximum intercuspation

The effects of MIC were examined using bilateral mean measurements for each muscle and mechanical property. Separate linear mixed-effects models were fitted for the SCM and trapezius muscles across the five myotonometric outcomes, yielding 10 models. Group (CG and NPG), experimental condition (rest and maximum intercuspation), and their interaction were specified as fixed effects, with a participant-specific random intercept to account for repeated observations. The use of bilateral mean measurements reflected the objective of examining the overall muscular response rather than side-specific differences.

Type III F-tests with Satterthwaite approximations to the denominator degrees of freedom were used to evaluate fixed effects. Model-based within-group contrasts estimated changes from rest to maximum intercuspation, with corresponding standard errors, 95% CIs, and p-values. Model assumptions were assessed through visual inspection of quantile–quantile plots and residual-versus-fitted plots, together with examination of residual dispersion across model strata.

Sensitivity analyses examined the stability of between-group differences in condition-related changes. Individual change scores were calculated as MIC minus rest. Between-group differences in mean change were estimated using 10,000 participant-level bootstrap resamples stratified by group, with percentile-based 95% CIs. Resampling was performed at the participant level to preserve the paired structure of the measurements used to calculate individual changes. Yuen’s independent-samples robust test, based on 20% trimmed means, was additionally applied to compare change scores between groups. The Holm step-down procedure was used to adjust p-values from these robust comparisons across the 10 myotonometric outcomes. These analyses were interpreted as complementary assessments of the stability of the between-group findings rather than as replacements for the primary mixed-effects models.

#### Exploratory between-group comparisons and correlation analyses

Exploratory analyses were conducted in a subsample of 36 participants, comprising 18 healthy controls and 18 individuals with chronic neck pain. Between-group differences were examined for six cervical range-of-motion measurements (flexion, extension, right and left rotation, and right and left lateral flexion), tongue pressure at 50%, 80%, and 100% of MTP, and five myotonometric properties (frequency, stiffness, logarithmic decrement, relaxation time, and creep). Myotonometric comparisons included bilateral summary measurements of the SCM and trapezius muscles obtained at rest and during the three tongue-pressure conditions.

Welch’s independent-samples t-test was used for exploratory between-group comparisons, allowing for unequal variances. Degrees of freedom were estimated using the Welch–Satterthwaite approximation. Mean differences were calculated as CG minus NPG and reported with 95% CIs. Hedges’ g was used to quantify standardized between-group differences, with positive values indicating higher measurements in the CG. Each variable and experimental condition was analyzed separately. These comparisons were considered exploratory, and their unadjusted p-values were interpreted accordingly.

Spearman’s rank-order correlation coefficient (ρ) was used to examine associations among cervical range of motion, tongue pressure, and myotonometric properties. Correlation analyses were performed separately for the CG and NPG, with 18 participants in each group. Myotonometric variables were represented by bilateral summary measurements of the SCM and trapezius muscles. Analyses were restricted to measurements obtained during the 50% and 100% maximum tongue-pressure conditions.

Within each group, correlations were examined between cervical range-of-motion measurements and myotonometric properties, between cervical range of motion and tongue pressure, and between tongue pressure and myotonometric properties. Pain intensity and neck disability were additionally included in the NPG analyses to examine their associations with the experimental measurements.

Spearman’s coefficients were calculated using complete observations for each variable pair. Statistical significance was initially evaluated using two-sided tests based on the asymptotic approximation. Correlation coefficients were displayed in heatmaps, with values rounded to two decimal places. Given the exploratory nature of these analyses and the number of correlations examined, the magnitude, direction, and clinical relevance of the coefficients were considered alongside their statistical significance.

As a supplementary sensitivity analysis, the Benjamini–Hochberg false discovery rate (FDR) procedure was applied to the Spearman correlation p-values to account for multiple testing. The adjustment was performed separately within the CG (132 correlations) and the neck pain group (176 correlations), treating each group as a distinct family of exploratory hypotheses. Associations with FDR-adjusted p-values below .05 were identified, while the original Spearman correlation coefficients and unadjusted p-values were retained. This procedure was used to examine the sensitivity of the exploratory findings to multiplicity rather than to replace the original correlation analyses.

#### Statistical significance, effect sizes, and software

All statistical tests were two-sided, with statistical significance set at p < .05. Adjustments for multiple comparisons were applied only where explicitly indicated. Omnibus p-values across separately fitted mixed-effects models and p-values from exploratory between-group comparisons were not adjusted. The supplementary FDR analysis of Spearman correlations was considered separately from the original exploratory inference.

Effect sizes were reported according to the statistical procedure: Hedges’ g for standardized mean differences, including Hedges’ g_av for within-participant changes; partial eta squared (η²p) for fixed effects in the tongue-pressure mixed-effects models; Cramér’s V for categorical associations; and Spearman’s ρ for rank-based correlations. For the MIC analyses, model-estimated changes and between-group differences in change were reported in their original measurement units with 95% CIs.

All analyses were performed using R statistical software (R Foundation for Statistical Computing, Vienna, Austria). Linear mixed-effects models and associated inferential procedures were implemented using the nlme, lme4, and lmerTest packages, as applicable. Estimated marginal means and post hoc contrasts were calculated using emmeans. Graphical representations were produced using ggplot2, and correlation matrices were visualized using corrplot. The Benjamini– Hochberg adjustment was implemented using the p.adjust function in R.

## 3. RESULTS AND DISCUSSION

### 3.1 RESULTS

#### 3.1.1 Participant characteristics

A total of 84 participants were included in the study, comprising 42 healthy controls (CG) and 42 individuals with chronic neck pain (NPG). Demographic and clinical characteristics are presented in Table 1.

**Table 1.** Demographic and clinical characteristics of the study population.

| **Characteristic** | **CG (n = 42)** | **NPG (n = 42)** | **Effect size** | **p** |
|---|---|---|---|---|
| Age (years) | 38.62 ± 16.10 | 39.02 ± 15.00 | −0.025 | .905 |
| **Sex, n (%)** |  |  | **0.229** | **.035** |
| Male | 18 (42.9) | 9 (21.4) |  |  |
| Female | 24 (57.1) | 33 (78.6) |  |  |
| **Employment status, n (%)** |  |  | 0.256 | **.240** |
| Student | 9 (21.4) | 6 (14.3) |  |  |
| Salaried employee | 20 (47.6) | 28 (66.7) |  |  |
| Self-employed | 5 (11.9) | 5 (11.9) |  |  |
| Unemployed | 6 (14.3) | 1 (2.4) |  |  |
| On sick leave | 2 (4.8) | 2 (4.8) |  |  |
| **Marital status, n (%)** |  |  | 0.118 | **.929** |
| Single | 26 (61.9) | 25 (59.5) |  |  |
| Married | 13 (31.0) | 15 (35.7) |  |  |
| Divorced | 2 (4.8) | 2 (4.8) |  |  |
| Widowed | 1 (2.4) | 0 (0.0) |  |  |
| **Clinical characteristics** |  |  |  |  |
| Pain intensity (NRS) | — | 5.75 ± 1.38 | — | — |
| Neck Disability Index (NDI) | — | 14.69 ± 6.00 | — | — |
| **Tongue pressure (kPa)** |  |  |  |  |
| 50% of maximum | 26.43 ± 5.15 | 24.14 ± 7.04 | 0.368 | .094 |
| 80% of maximum | 42.40 ± 8.21 | 37.47 ± 9.46 | 0.552 | .013 |
| 100% of maximum | 52.99 ± 10.26 | 47.33 ± 12.01 | 0.502 | .023 |

**Note.** Values are mean ± standard deviation or n (%). CG = control group; NPG = chronic neck pain group; NRS = numerical rating scale; NDI = Neck Disability Index.
Effect sizes are Hedges’ g for continuous variables and Cramér’s V for sex, employment status, and marital status. Hedges’ g was calculated using the pooled standard deviation and small-sample correction, with CG − NPG as the direction of comparison. Cramér’s V was calculated from the Pearson chi-square statistic and the corresponding contingency-table dimensions.
Continuous variables were compared using Welch’s independent-samples t-test; sex using Pearson’s chi-square test; employment and marital status using Fisher’s exact test. All p-values are unadjusted. — = not applicable or not calculated.

Mean age was 38.62 ± 16.10 years in the CG and 39.02 ± 15.00 years in the NPG (*p* = .905). Women accounted for 57.1% (n = 24) of the CG and 78.6% (n = 33) of the NPG (χ²(1) = 4.42, *p* = .035). Between-group comparisons of employment status (*p* = .240) and marital status (*p* = .929) were also performed. In the NPG, mean pain intensity was 5.75 ± 1.38 points on the NRS, and the mean NDI score was 14.69 ± 6.00 points.

At 50% of MTP, mean values were 26.43 ± 5.15 kPa in the CG and 24.14 ± 7.04 kPa in the NPG (mean difference [CG − NPG] = 2.28 kPa, 95% CI [−0.40, 4.97], *p* = .094). At 80% MTP, mean tongue pressure was 42.40 ± 8.21 kPa in the CG and 37.47 ± 9.46 kPa in the NPG (mean difference = 4.92 kPa, 95% CI [1.08, 8.77], *p* = .013). At 100% MTP, the corresponding values were 52.99 ± 10.26 and 47.33 ± 12.01 kPa (mean difference = 5.66 kPa, 95% CI [0.81, 10.51], *p* = .023).

#### 3.1.2 Preliminary data screening

Preliminary screening of the myotonometric data included the examination of distributional characteristics, potential outliers, and between-group variance homogeneity. Brown–Forsythe tests were performed for 125 combinations of experimental condition, anatomical location, and mechanical property. Ten comparisons (8.0%) yielded unadjusted p-values below .05, comprising four comparisons for stiffness, three for muscle frequency, two for relaxation time, and one for creep. No comparisons involving logarithmic decrement yielded unadjusted p-values below .05. None of the 125 comparisons remained statistically significant after Benjamini–Hochberg adjustment.

The data audit identified a transcription error in a right trapezius stiffness measurement obtained at 80% MTP in a CG participant. Following verification against the original measurement record, the recorded value was corrected from 2667 to 267 N/m. The corresponding corrected mean of the two measurements was 267.5 N/m.

Visual inspection of quantile–quantile and residual-versus-fitted plots for the 20 tongue-pressure mixed-effects models identified deviations from residual normality in the distribution tails and differences in residual dispersion in selected models.

#### 3.1.3. Muscle frequency

A significant main effect of condition was observed for muscle frequency at all four anatomical locations (all *p* < .001; partial η² = .073–.119). Neither the main effect of group nor the group-by-condition interaction was statistically significant at any location (all *p* > .05; Table 2).

**Table 2.**
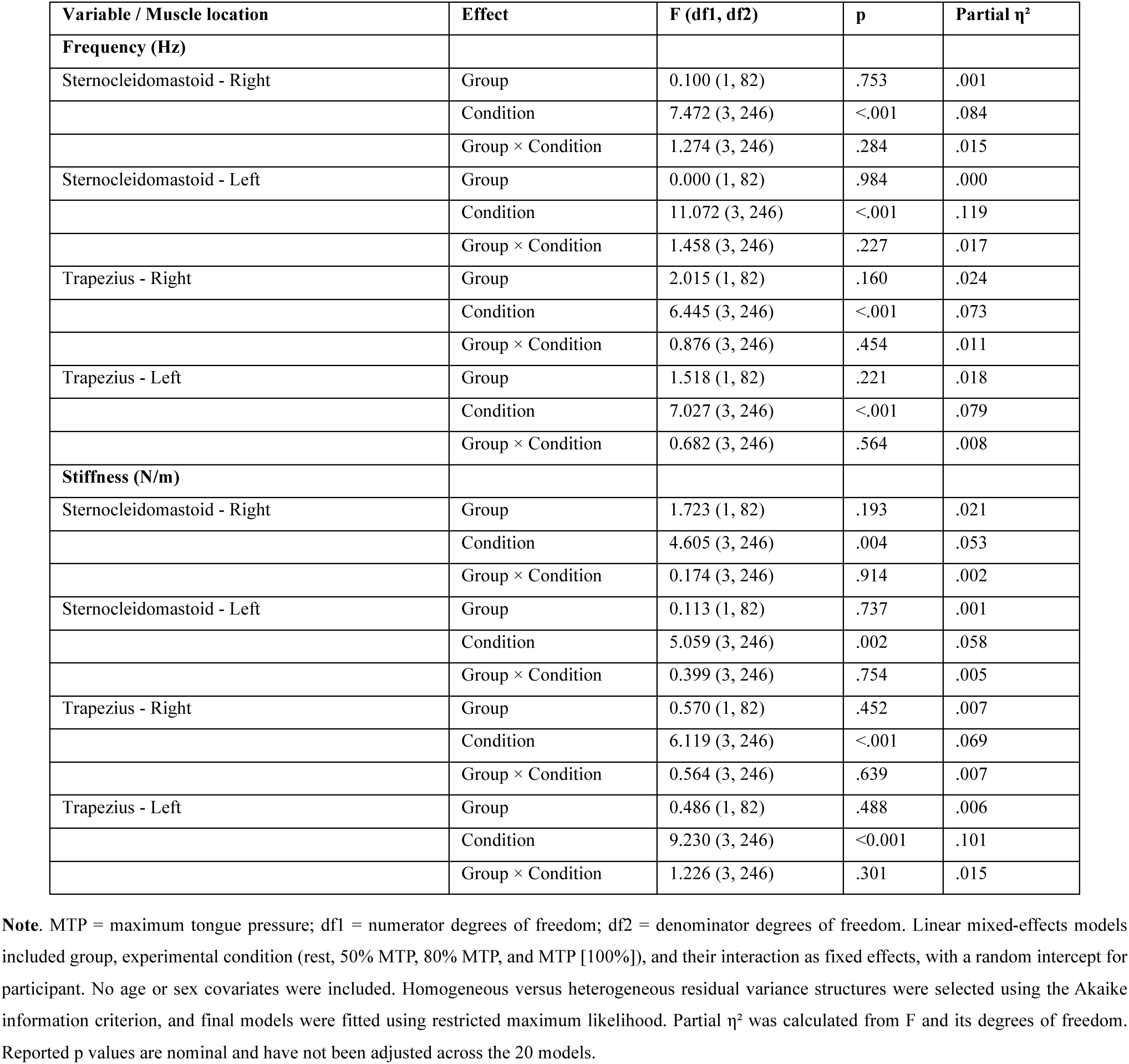
Global effects of group and experimental condition on muscle frequency and stiffness.

Post hoc comparisons showed significant increases in frequency relative to rest in the NPG at 80% and 100% MTP in the right SCM (Hedges’ *g* = 0.41 and 0.39, respectively) and left SCM (*g* = 0.37 and 0.55, respectively). No significant changes relative to rest were identified in the CG at either SCM location.

In the right trapezius, significant increases were observed in NPG at 50% and 100% MTP (*g* = 0.32 and 0.31, respectively), whereas CG showed a significant increase at 100% MTP (*g* = 0.35). In the left trapezius, significant increases were identified in NPG at 50%, 80%, and 100% MTP (*g* = 0.25, 0.25, and 0.29, respectively). The CG also exhibited a significant increase at 80% MTP (*g* = 0.36).

Despite these within-group changes, no significant differences in muscle frequency were identified between CG and NPG at any experimental condition. The complete within-group and between-group comparisons are presented in Supplementary Table S1, and the corresponding trajectories are illustrated in Figure 1.

**Figure 1.**
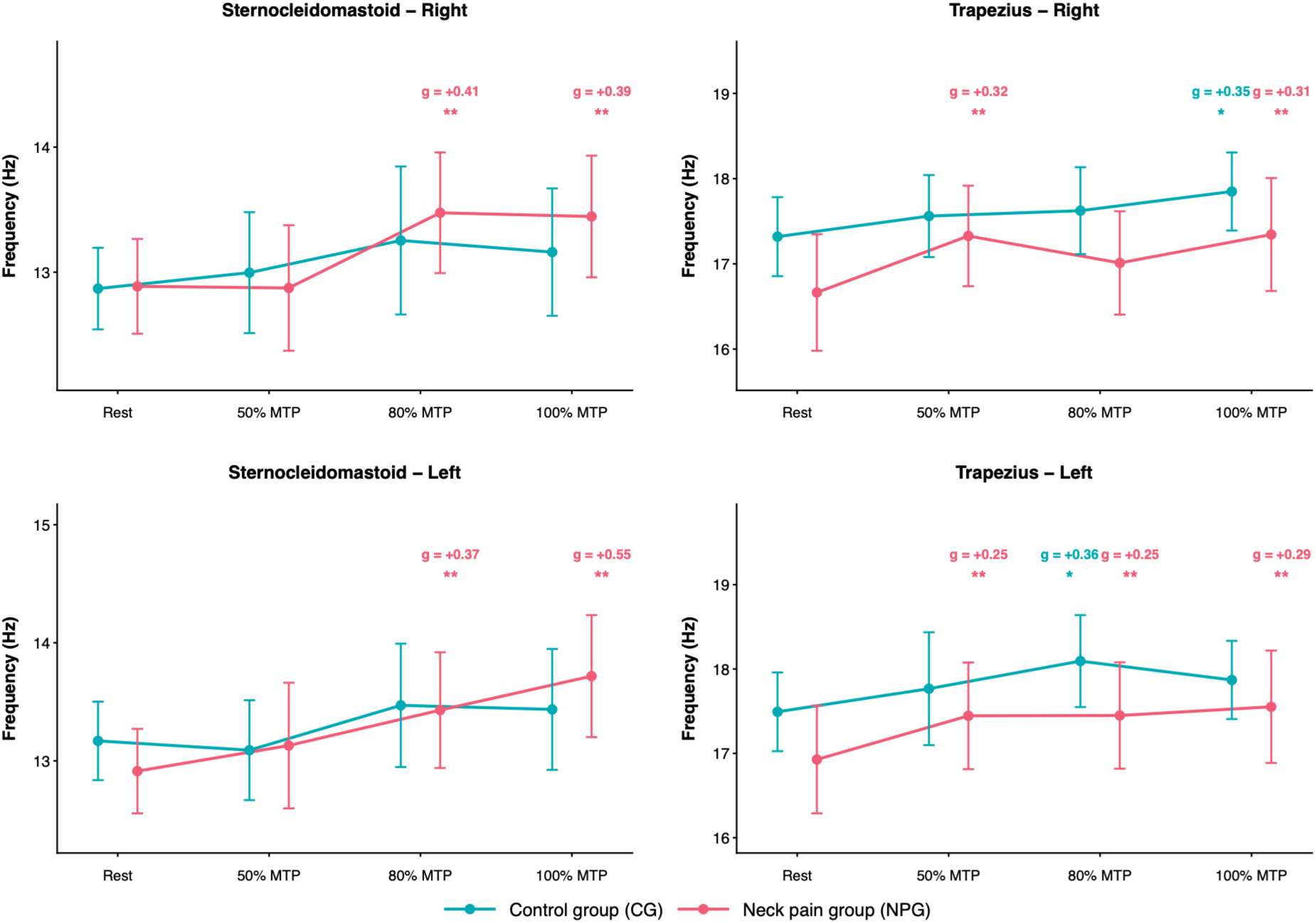
Muscle frequency across tongue-pressure conditions in healthy controls and participants with chronic neck pain. Mean muscle oscillation frequency (Hz) in the right and left sternocleidomastoid and trapezius muscles at rest and during tongue-pressure tasks performed at 50%, 80%, and 100% of maximum tongue pressure (MTP). The control group (CG; n = 42) and neck pain group (NPG; n = 42) are represented separately. Points indicate observed group means, and error bars represent descriptive 95% confidence intervals. Asterisks indicate statistically significant within-group differences relative to rest based on Holm-adjusted post hoc comparisons (p < .05 [*] and p < .01 [**]). These markers do not represent between-group differences. Hedges’ g values, where displayed, describe the standardized magnitude of the corresponding within-group contrasts. Statistical annotations refer exclusively to comparisons with the resting condition and should not be interpreted as evidence of between-group differences.

#### 3.1.4. Muscle stiffness

A statistically significant main effect of experimental condition on muscle stiffness was identified at all four anatomical locations (p ≤ .004; partial η² = .053–.101). Neither the main effect of group nor the group-by-condition interaction reached statistical significance at any location (all p > .05; Table 2).

Post hoc comparisons identified significant increases in stiffness relative to rest in the NPG at 80% and 100% MTP in the right SCM (Hedges’ g = 0.28 and 0.27, respectively) and left SCM (g = 0.24 and 0.36, respectively). No significant changes relative to rest were identified in either SCM muscle in the CG.

In the right trapezius, stiffness increased significantly in the NPG at 50%, 80%, and 100% MTP (g = 0.31, 0.18, and 0.27, respectively), whereas the CG showed a significant increase at 100% MTP (g = 0.32). In the left trapezius, significant increases were identified in the NPG at 50%, 80%, and 100% MTP (g = 0.30, 0.29, and 0.37, respectively), while the CG showed a significant increase at 80% MTP (g = 0.36).

No statistically significant between-group differences in muscle stiffness were identified at any anatomical location or experimental condition. Detailed post hoc comparisons are presented in Supplementary Table S2, and the corresponding measurements across tongue-pressure conditions are illustrated in Figure 2.

**Figure 2.**
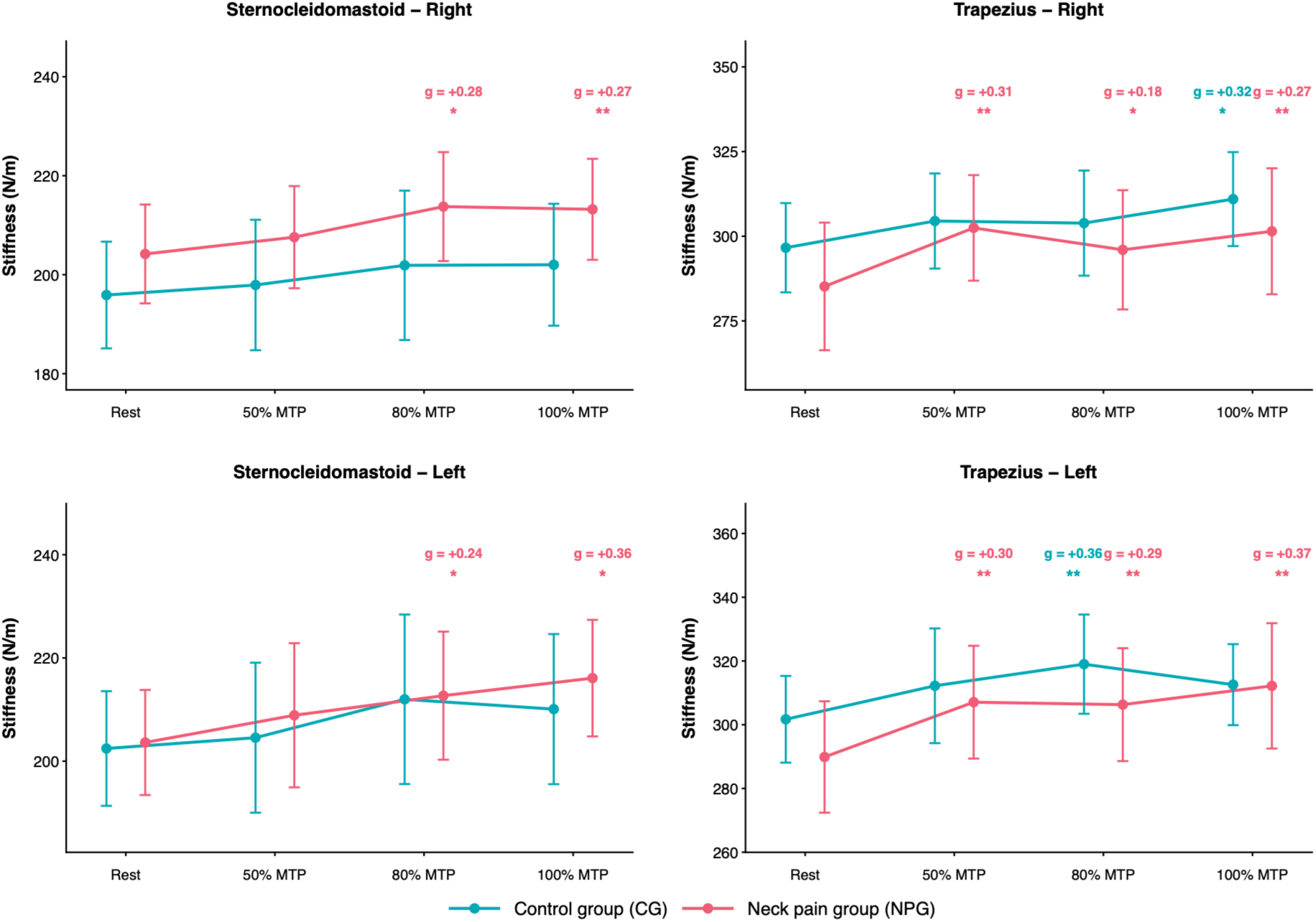
Muscle stiffness across tongue-pressure conditions in healthy controls and participants with chronic neck pain. Mean muscle stiffness (N/m) measured bilaterally in the sternocleidomastoid and trapezius muscles under four experimental conditions: rest and tongue-pressure tasks at 50%, 80%, and 100% of maximum tongue pressure (MTP). Data are presented for the control group (CG; n = 42) and neck pain group (NPG; n = 42). Points represent observed means, with error bars indicating descriptive 95% confidence intervals. Asterisks indicate statistically significant within-group differences relative to rest based on Holm-adjusted post hoc comparisons (p < .05 [*] and p < .01 [**]). These markers do not represent between-group differences. Hedges’ g values, where shown, quantify the standardized magnitude of these contrasts. The significance markers do not represent direct comparisons between groups.

#### 3.1.5. Muscle relaxation time

A statistically significant main effect of experimental condition on muscle relaxation time was identified at all four anatomical locations (p ≤ .008; partial η² = .047–.114). Neither the main effect of group nor the group-by-condition interaction reached statistical significance at any location (all p > .05; Table 3).

**Table 3.**
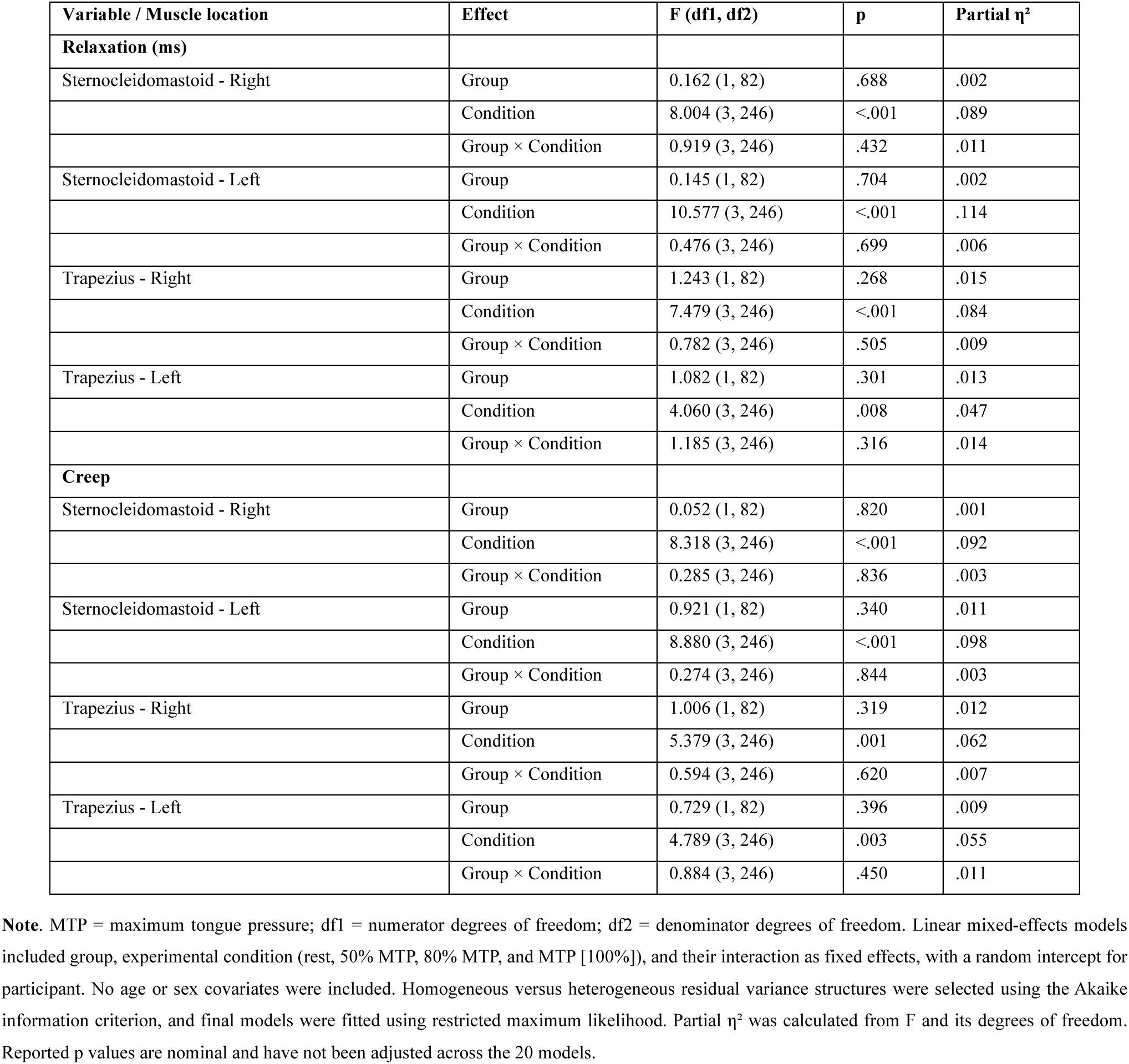
Global effects of group and experimental condition on muscle relaxation and creep.

| **Variable / Muscle location** | **Effect** | **F (df1, df2)** | **p** | **Partial η²** |
|---|---|---|---|---|
| **Relaxation (ms)** |  |  |  |  |
| Sternocleidomastoid - Right | Group | 0.162 (1, 82) | .688 | .002 |
|  | Condition | 8.004 (3, 246) | <.001 | .089 |
|  | Group × Condition | 0.919 (3, 246) | .432 | .011 |
| Sternocleidomastoid - Left | Group | 0.145 (1, 82) | .704 | .002 |
|  | Condition | 10.577 (3, 246) | <.001 | .114 |
|  | Group × Condition | 0.476 (3, 246) | .699 | .006 |
| Trapezius - Right | Group | 1.243 (1, 82) | .268 | .015 |
|  | Condition | 7.479 (3, 246) | <.001 | .084 |
|  | Group × Condition | 0.782 (3, 246) | .505 | .009 |
| Trapezius - Left | Group | 1.082 (1, 82) | .301 | .013 |
|  | Condition | 4.060 (3, 246) | .008 | .047 |
|  | Group × Condition | 1.185 (3, 246) | .316 | .014 |
| **Creep** |  |  |  |  |
| Sternocleidomastoid - Right | Group | 0.052 (1, 82) | .820 | .001 |
|  | Condition | 8.318 (3, 246) | <.001 | .092 |
|  | Group × Condition | 0.285 (3, 246) | .836 | .003 |
| Sternocleidomastoid - Left | Group | 0.921 (1, 82) | .340 | .011 |
|  | Condition | 8.880 (3, 246) | <.001 | .098 |
|  | Group × Condition | 0.274 (3, 246) | .844 | .003 |
| Trapezius - Right | Group | 1.006 (1, 82) | .319 | .012 |
|  | Condition | 5.379 (3, 246) | .001 | .062 |
|  | Group × Condition | 0.594 (3, 246) | .620 | .007 |
| Trapezius - Left | Group | 0.729 (1, 82) | .396 | .009 |
|  | Condition | 4.789 (3, 246) | .003 | .055 |
|  | Group × Condition | 0.884 (3, 246) | .450 | .011 |

**Note**. MTP = maximum tongue pressure; df1 = numerator degrees of freedom; df2 = denominator degrees of freedom. Linear mixed-effects models included group, experimental condition (rest, 50% MTP, 80% MTP, and MTP [100%]), and their interaction as fixed effects, with a random intercept for participant. No age or sex covariates were included. Homogeneous versus heterogeneous residual variance structures were selected using the Akaike information criterion, and final models were fitted using restricted maximum likelihood. Partial η² was calculated from F and its degrees of freedom. Reported p values are nominal and have not been adjusted across the 20 models.

Post hoc comparisons identified significant reductions in relaxation time relative to rest in the NPG at 80% and 100% of MTP in the right SCM (Hedges’ g = −0.43 and −0.36, respectively) and left SCM (g = −0.29 and −0.39, respectively). No significant changes relative to rest were identified in either SCM muscle in the CG.

In the right trapezius, relaxation time decreased significantly in the NPG at 50%, 80%, and 100% MTP (g = −0.35, −0.21, and −0.37, respectively), whereas the CG showed a significant reduction at 100% MTP (g = −0.41). In the left trapezius, significant reductions were identified in the NPG at 50%, 80%, and 100% MTP (g = −0.26, −0.25, and −0.33, respectively). No significant changes relative to rest were identified in the left trapezius of the CG.

No statistically significant between-group differences in relaxation time were identified at any anatomical location or experimental condition. The complete post hoc comparisons are presented in Supplementary Table S3, and the condition-specific measurements are illustrated in Figure 3.

**Figure 3.**
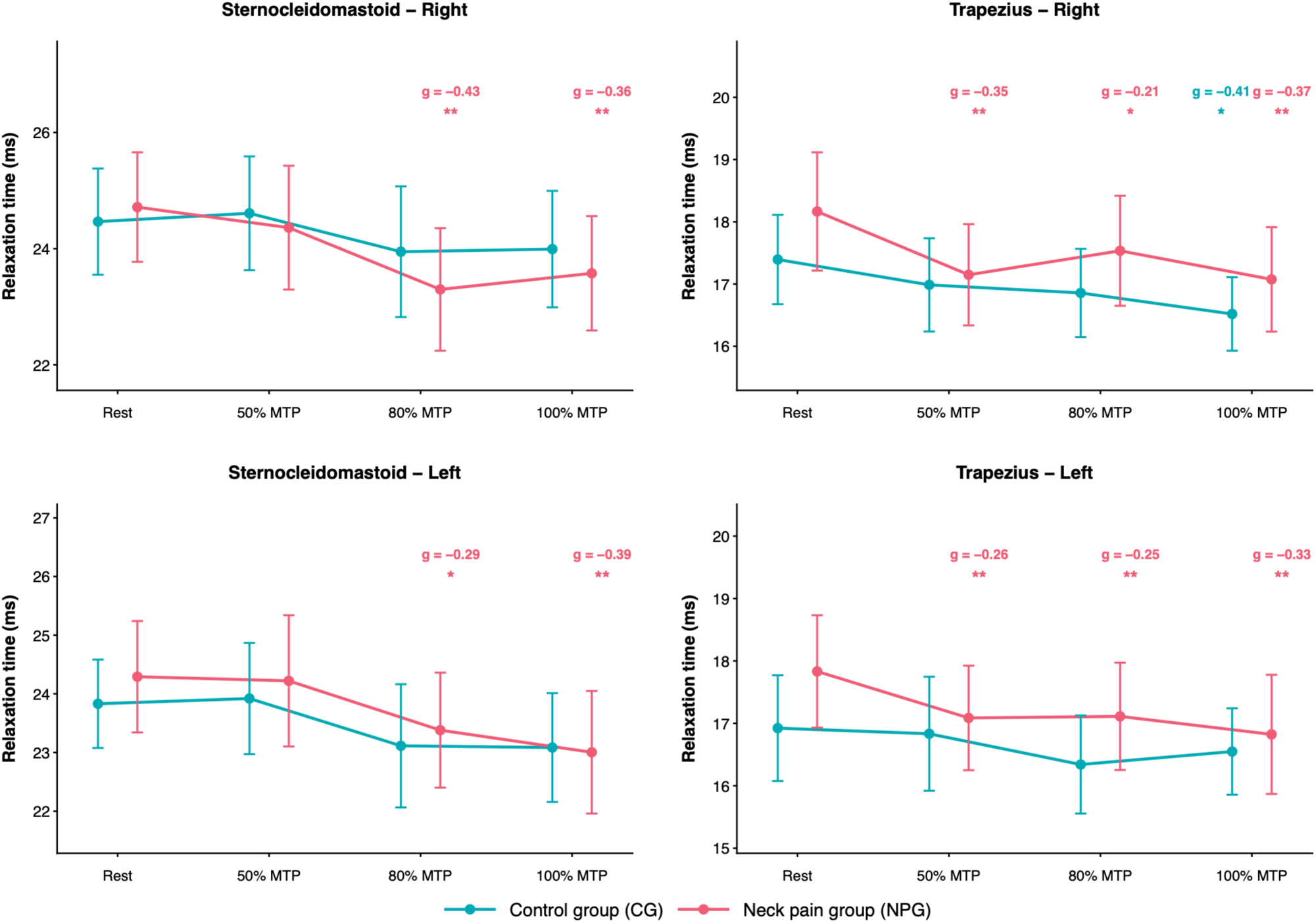
Muscle relaxation time across tongue-pressure conditions in healthy controls and participants with chronic neck pain. Mean muscle relaxation time (ms) in the right and left sternocleidomastoid and trapezius muscles during rest and tongue-pressure tasks performed at 50%, 80%, and 100% of maximum tongue pressure (MTP). Results are displayed separately for the control group (CG; n = 42) and neck pain group (NPG; n = 42). Points represent observed group means, and error bars indicate descriptive 95% confidence intervals. Asterisks indicate statistically significant within-group differences relative to rest based on Holm-adjusted post hoc comparisons (p < .05 [*] and p < .01 [**]). These markers do not represent between-group differences. Hedges’ g values, where provided, indicate the standardized magnitude and direction of the corresponding contrasts. Negative effect-size values indicate lower relaxation time during the tongue-pressure condition relative to rest. The annotations do not indicate statistically significant between-group differences.

#### 3.1.6. Muscle creep

A statistically significant main effect of experimental condition on muscle creep was identified at all four anatomical locations (p ≤ .003; partial η² = .055–.098). Neither the main effect of group nor the group-by-condition interaction reached statistical significance at any location (all p > .05; Table 3).

Post hoc comparisons identified significant reductions in creep relative to rest in the NPG at 80% and 100% of MTP in the right SCM (Hedges’ g = −0.40 and −0.31, respectively). In the left SCM, a significant reduction was identified at 100% MTP (g = −0.31). No significant changes relative to rest were identified in either SCM muscle in the CG.

In the right trapezius, creep decreased significantly in the NPG at 50% and 100% MTP (g = −0.29 and −0.38, respectively), whereas the CG showed a significant reduction at 100% MTP (g = −0.40). In the left trapezius, a significant reduction was identified in the NPG at 100% MTP (g = −0.33), with no significant changes relative to rest in the CG.

No statistically significant between-group differences in creep were identified at any anatomical location or experimental condition. Detailed post hoc comparisons are presented in Supplementary Table S4, and the corresponding measurements across tongue-pressure conditions are illustrated in Figure 4.

**Figure 4.**
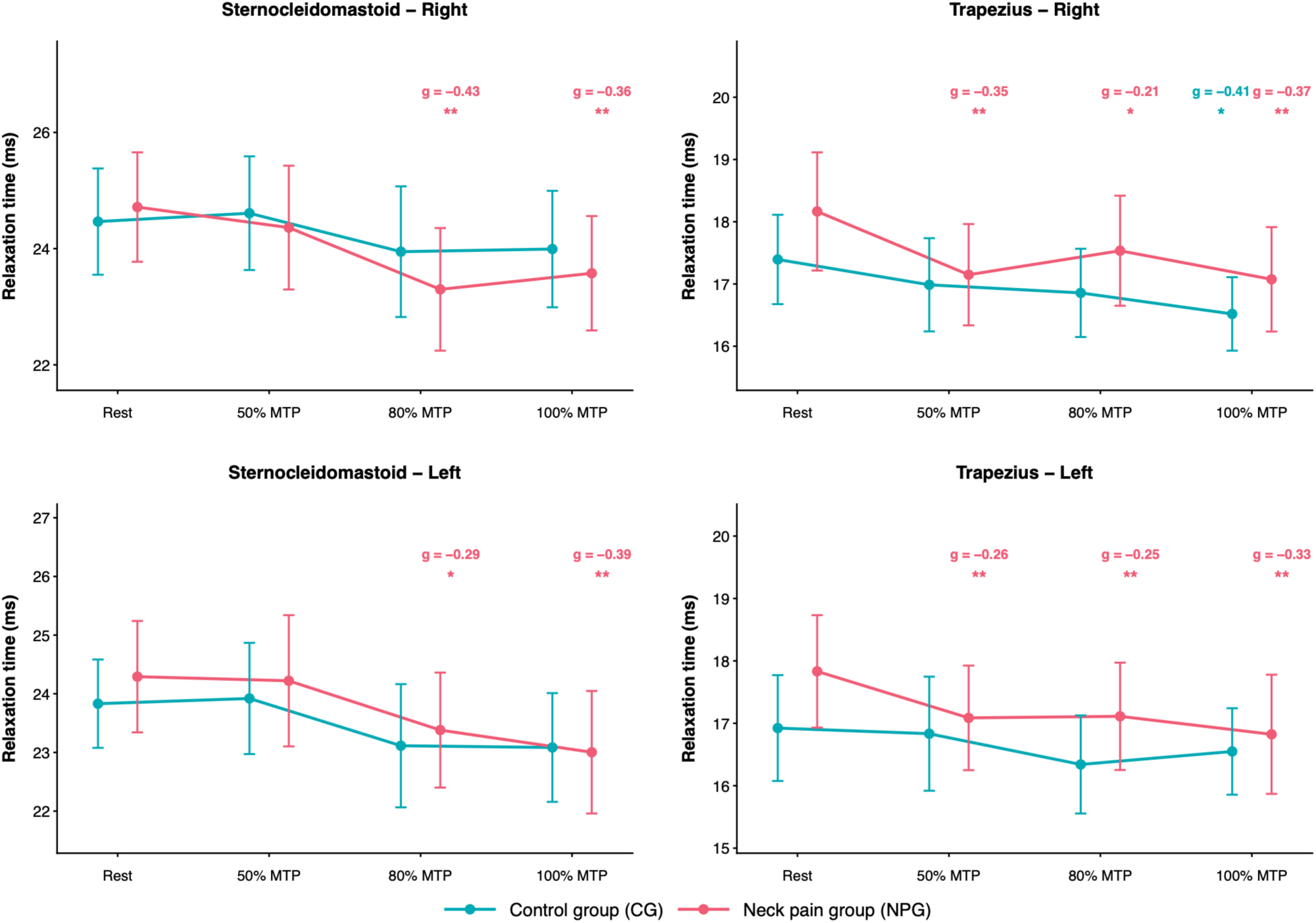
Muscle creep across tongue-pressure conditions in healthy controls and participants with chronic neck pain. Mean muscle creep (dimensionless) recorded bilaterally in the sternocleidomastoid and trapezius muscles at rest and during tongue-pressure tasks at 50%, 80%, and 100% of maximum tongue pressure (MTP). The control group (CG; n = 42) and neck pain group (NPG; n = 42) are displayed separately. Points indicate observed means, and error bars represent descriptive 95% confidence intervals. Asterisks indicate statistically significant within-group differences relative to rest based on Holm-adjusted post hoc comparisons (p < .05 [*] and p < .01 [**]). These markers do not represent between-group differences. Hedges’ g values, where displayed, represent the standardized magnitude and direction of the corresponding contrasts. Negative values indicate reduced creep during the tongue-pressure task compared with rest. The statistical annotations are restricted to within-group comparisons and do not establish differences between groups.

#### 3.1.7. Muscle decrement

Statistically significant main effects of experimental condition on logarithmic decrement were identified in the right SCM (F(3, 246) = 3.225, p = .023, partial η² = .038), right trapezius (F(3, 246) = 3.057, p = .029, partial η² = .036), and left trapezius (F(3, 246) = 5.828, p < .001, partial η² = .066). The main effect of condition was not statistically significant in the left SCM (F(3, 246) = 1.355, p = .257, partial η² = .016; Table 4).

**Table 4.** Global effects of group and experimental condition on muscle decrement.

| **Variable / Muscle location** | **Effect** | **F (df1, df2)** | **p** | **Partial η²** |
|---|---|---|---|---|
| **Decrement** |  |  |  |  |
| Sternocleidomastoid - Right | Group | 1.129 (1, 82) | .291 | .014 |
|  | Condition | 3.225 (3, 246) | .023 | .038 |
|  | Group × Condition | 0.764 (3, 246) | .515 | .009 |
| Sternocleidomastoid - Left | Group | 0.268 (1, 82) | .606 | .003 |
|  | Condition | 1.355 (3, 246) | .257 | .016 |
|  | Group × Condition | 2.274 (3, 246) | .081 | .027 |
| Trapezius - Right | Group | 1.360 (1, 82) | .247 | .016 |
|  | Condition | 3.057 (3, 246) | .029 | .036 |
|  | Group × Condition | 3.248 (3, 246) | .023 | .038 |
| Trapezius - Left | Group | 3.527 (1, 82) | .064 | .041 |
|  | Condition | 5.828 (3, 246) | <.001 | .066 |
|  | Group × Condition | 1.069 (3, 246) | .363 | .013 |

**Note**. MTP = maximum tongue pressure; df1 = numerator degrees of freedom; df2 = denominator degrees of freedom. Linear mixed-effects models included group, experimental condition (rest, 50% MTP, 80% MTP, and MTP [100%]), and their interaction as fixed effects, with a random intercept for participant. No age or sex covariates were included. Homogeneous versus heterogeneous residual variance structures were selected using the Akaike information criterion, and final models were fitted using restricted maximum likelihood. Partial η² was calculated from F and its degrees of freedom. Reported p values are nominal and have not been adjusted across the 20 models.

No statistically significant main effects of group were identified at any anatomical location (all p > .05). A statistically significant group-by-condition interaction was observed for right trapezius logarithmic decrement (F(3, 246) = 3.248, p = .023, partial η² = .038). The corresponding interactions were not statistically significant in the right SCM (p = .515), left SCM (p = .081), or left trapezius (p = .363).

Within-group post hoc comparisons identified statistically significant increases in logarithmic decrement relative to rest in the NPG at 50% and 80% of MTP in the right trapezius (Hedges’ g_av = 0.25 and 0.27, respectively). In the left trapezius, significant increases were identified at 50%, 80%, and 100% MTP (g_av = 0.38, 0.36, and 0.27, respectively). No statistically significant changes relative to rest were identified in either SCM muscle or in the trapezius muscles of the CG. These within-group findings describe condition-related changes within each group and do not, by themselves, establish differences in the magnitude of change between groups. Complete within-group contrasts are presented in Supplementary Table S5A.

Between-group comparisons did not identify statistically significant differences in logarithmic decrement at any anatomical location or experimental condition after Holm adjustment. In the left trapezius at 80% MTP, the estimated difference between groups was −0.076 (95% CI [−0.147, −0.005]); however, this comparison did not remain statistically significant after adjustment for multiple comparisons (Holm-adjusted p = .145). The confidence interval was not adjusted for multiplicity and should therefore be interpreted separately from the adjusted p value. Complete condition-specific between-group comparisons are presented in Supplementary Table S5B.

Although a statistically significant group-by-condition interaction was identified for right trapezius logarithmic decrement, the corresponding between-group differences in changes from rest were not statistically significant after Holm adjustment. The estimated differences (NPG − CG) were 0.043 at 50% MTP (95% CI [−0.011, 0.098], adjusted p = .114), 0.044 at 80% MTP (95% CI [−0.006, 0.094], adjusted p = .105), and −0.001 at 100% MTP (95% CI [−0.060, 0.057], adjusted p = .965). Thus, the significant omnibus interaction was not accompanied by statistically significant individual difference-in-change contrasts. Complete condition-specific between-group comparisons are presented in Supplementary Table S5B, and the corresponding measurements are illustrated in Figure 5.

**Figure 5.**
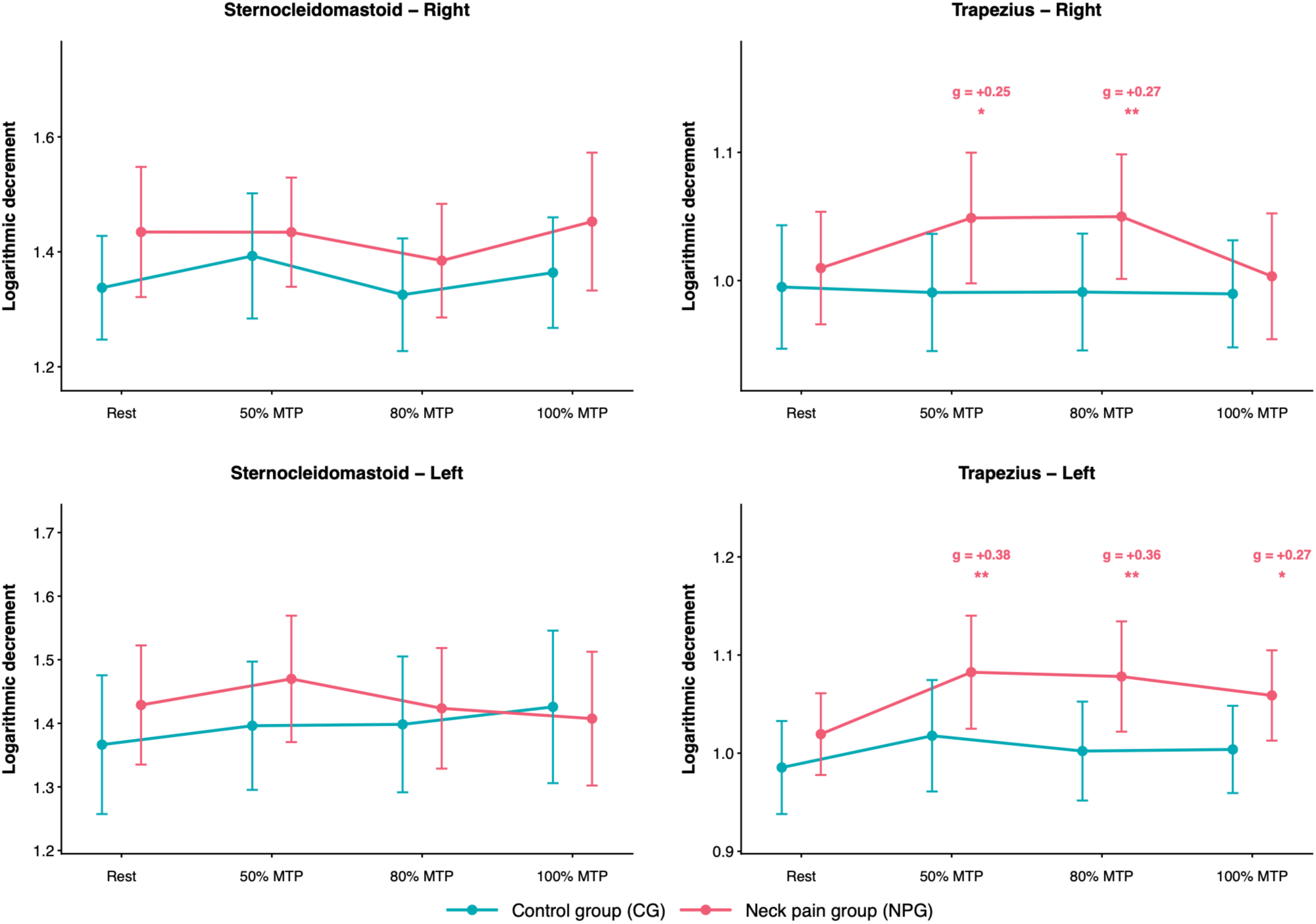
Muscle decrement across tongue-pressure conditions in healthy controls and participants with chronic neck pain. Mean logarithmic decrement (dimensionless) in the right and left sternocleidomastoid and trapezius muscles at rest and during tongue-pressure tasks performed at 50%, 80%, and 100% of maximum tongue pressure (MTP). Results are presented separately for the control group (CG; n = 42) and neck pain group (NPG; n = 42). Points represent observed means, and error bars indicate descriptive 95% confidence intervals. Asterisks indicate statistically significant within-group differences relative to rest based on Holm-adjusted post hoc comparisons (p < .05 [*] and p < .01 [**]). These markers do not represent between-group differences. Hedges’ g values, where displayed, describe the standardized magnitude of these contrasts. The mixed-effects analysis identified a group-by-condition interaction for right trapezius logarithmic decrement (F(3, 246) = 3.248, p = .0225, partial η² = .038). This interaction reflects differences in the pattern of responses across experimental conditions and should be distinguished from the within-group comparisons indicated by the asterisks.

#### 3.1.8. Myotonometric responses to maximum intercuspation

The analysis of myotonometric responses to MIC included 84 participants, comprising 42 healthy controls (CG) and 42 individuals with NPG. Descriptive statistics and linear mixed-effects model results for the trapezius and SCM muscles are presented in Table 5. No statistically significant main effects of group were identified for any of the ten myotonometric outcomes (all p > .05).

**Table 5.** Descriptive statistics and mixed-model ANOVA for muscle mechanical properties according to group and experimental condition.

| Property | CG<br>Rest / MIC | NPG<br>Rest / MIC | Group<br>F (p) | Condition<br>F (p) | Group ×<br>Condition<br>F (p) |
|---|---|---|---|---|---|
| **Trapezius** |  |  |  |  |  |
| Frequency (Hz) | 17.41 ± 1.40 17.60 ± 1.42 | 16.80 ± 1.98 16.99 ± 2.03 | 2.76 (.101) | **4.17 (.044)** | 0.00 (.978) |
| Stiffness (N/m) | 299.15 ± 40.61 304.37 ± 43.33 | 287.53 ± 54.56 292.96 ± 56.74 | 1.21 (.275) | **4.66 (.034)** | 0.00 (.964) |
| Logarithmic Decrement | 0.99 ± 0.14 0.99 ± 0.13 | 1.01 ± 0.13 1.04 ± 0.14 | 1.48 (.227) | 3.92 (.051) | 3.12 (.081) |
| Relaxation time (ms) | 17.16 ± 2.31 17.03 ± 2.24 | 18.00 ± 2.69 17.73 ± 2.67 | 2.22 (.140) | 1.51 (.223) | 0.18 (.675) |
| Creep | 1.06 ± 0.13 1.05 ± 0.13 | 1.10 ± 0.15 1.08 ± 0.15 | 1.56 (.215) | 2.27 (.136) | 0.01 (.924) |
| **Sternocleidomastoid** |  |  |  |  |  |
| Frequency (Hz) | 13.02 ± 0.98 13.30 ± 1.26 | 12.90 ± 1.14 13.12 ± 1.02 | 0.43 (.515) | **9.20 (.003)** | 0.13 (.719) |
| Stiffness (N/m) | 199.17 ± 33.39 206.01 ± 36.17 | 203.90 ± 31.20 206.92 ± 27.10 | 0.17 (.678) | **7.11 (.009)** | 1.07 (.304) |
| Logarithmic Decrement | 1.35 ± 0.31 1.36 ± 0.33 | 1.43 ± 0.32 1.42 ± 0.31 | 1.02 (.315) | 0.09 (.766) | 0.66 (.417) |
| Relaxation time (ms) | 24.15 ± 2.47 23.56 ± 2.65 | 24.50 ± 2.85 24.03 ± 2.64 | 0.58 (.449) | **6.43 (.013)** | 0.07 (.791) |
| Creep | 1.41 ± 0.14 1.38 ± 0.16 | 1.45 ± 0.17 1.42 ± 0.17 | 1.52 (.222) | **5.19 (.025)** | 0.33 (.568) |

**Note.** Values are mean ± SD. In each group column, the first line represents Rest and the second line represents MIC. CG: healthy controls (n = 42); NPG: chronic neck pain (n = 42); MIC: maximum intercuspation. F statistics are Type III tests from linear mixed-effects models with Satterthwaite degrees of freedom (numerator df = 1; denominator df approximately 82). Unadjusted p values are reported; significant condition effects (p < .05) are bold. Bilateral muscle values were averaged.

In the trapezius, statistically significant main effects of experimental condition were identified for muscle frequency (F(1, 82) = 4.17, p = .044) and stiffness (F(1, 82) = 4.66, p = .034). Across groups, both properties showed higher mean values during MIC than at rest. The main effect of condition was not statistically significant for logarithmic decrement (F(1, 82) = 3.92, p = .051), relaxation time (F(1, 82) = 1.51, p = .223), or creep (F(1, 82) = 2.27, p = .136).

In the SCM, statistically significant main effects of condition were identified for frequency (F(1, 82) = 9.20, p = .003), stiffness (F(1, 82) = 7.11, p = .009), relaxation time (F(1, 82) = 6.43, p = .013), and creep (F(1, 82) = 5.19, p = .025). Across groups, mean frequency and stiffness were higher during MIC than at rest, whereas relaxation time and creep were lower. No statistically significant main effect of condition was identified for logarithmic decrement (F(1, 82) = 0.09, p = .766).

None of the group-by-condition interactions reached statistical significance across the ten outcomes examined (all p > .05). The lowest interaction p-value was observed for trapezius logarithmic decrement (F(1, 82) = 3.12, p = .081). The distributions of the five myotonometric properties under resting and MIC conditions are presented for the trapezius in Figure 6 and for the SCM in Figure 7.

**Figure 6.**
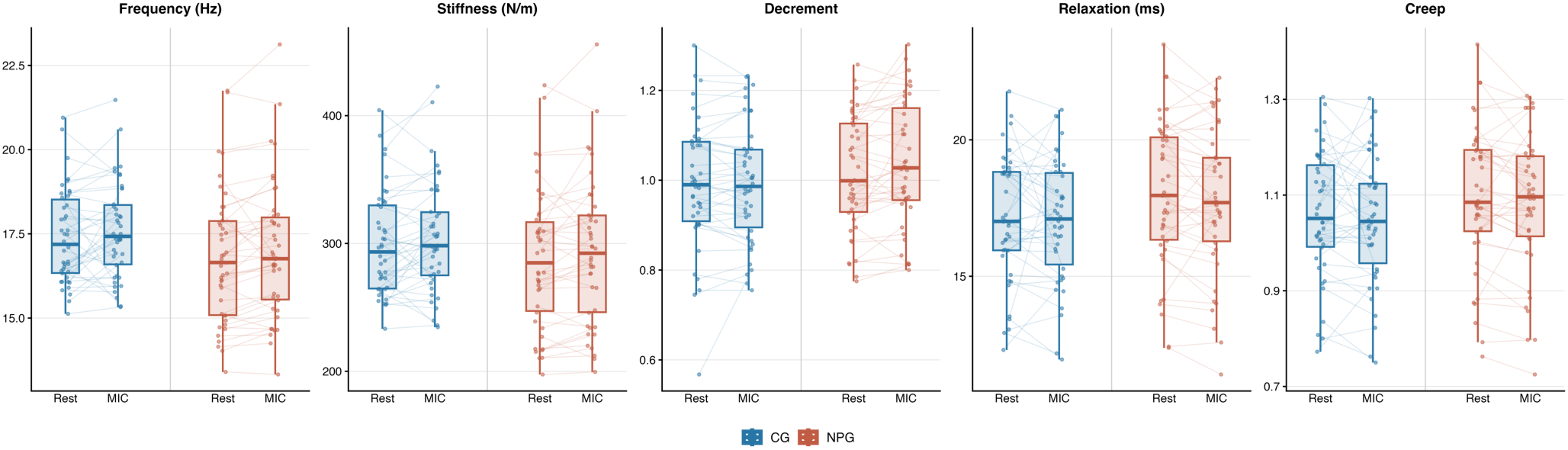
Distribution of trapezius mechanical properties at rest and during maximum intercuspation in healthy controls and participants with chronic neck pain. Box-and-whisker plots show the distributions of muscle frequency, stiffness, decrement, relaxation, and creep under resting (Rest) and MIC conditions. Boxes represent the interquartile range, horizontal lines indicate the median, and whiskers extend to the most extreme observations within 1.5 times the interquartile range. Individual points represent participant-level measurements, and connecting lines indicate paired observations across conditions. Blue represents healthy controls (CG; n = 42), and terracotta represents participants with chronic neck pain (NPG; n = 42). Measurements correspond to bilateral muscle averages.

**Figure 7.**
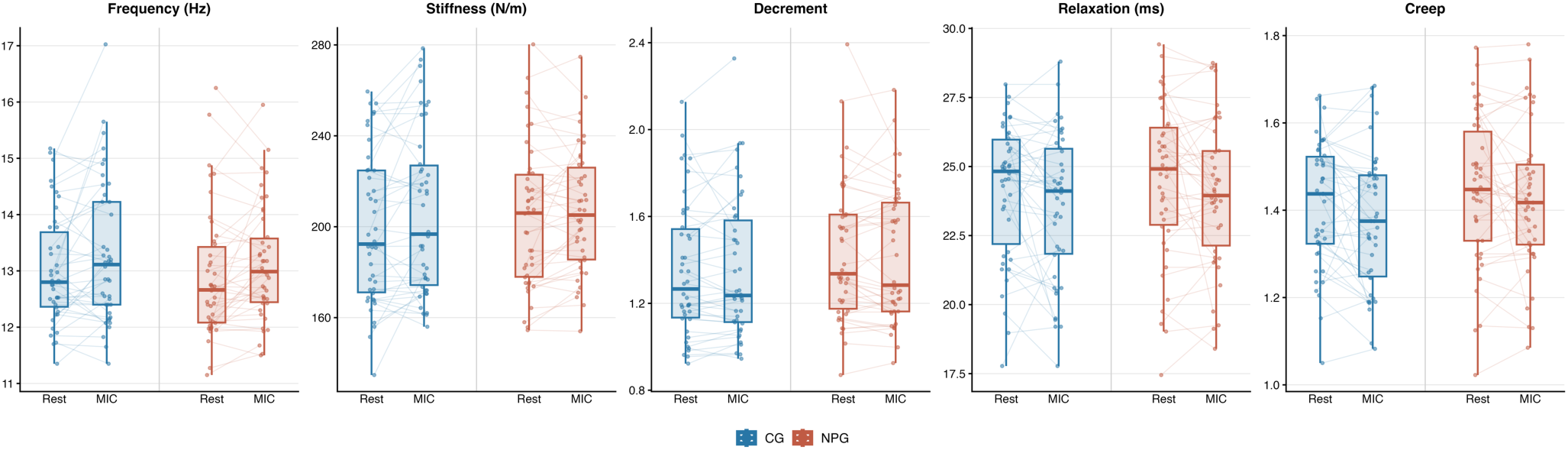
Distribution of sternocleidomastoid mechanical properties at rest and during maximum intercuspation in healthy controls and participants with chronic neck pain. Box-and-whisker plots show the distributions of muscle frequency, stiffness, decrement, relaxation, and creep under resting (Rest) and MIC conditions. Boxes represent the interquartile range, horizontal lines indicate the median, and whiskers extend to the most extreme observations within 1.5 times the interquartile range. Individual points represent participant-level measurements, and connecting lines indicate paired observations across conditions. Blue represents healthy controls (CG; n = 42), and terracotta represents participants with chronic neck pain (NPG; n = 42). Measurements correspond to bilateral muscle averages.

#### 3.1.9. Within-group changes during maximum intercuspation

Within-group contrasts were calculated to quantify changes in myotonometric properties from rest to MIC in the CG and NPG. Model-estimated mean differences, standard errors, 95% confidence intervals, and unadjusted p-values are presented in Table 6.

**Table 6.** Within-group changes in muscle mechanical properties from rest to maximum intercuspation.

| **Muscle** | **Property** | **Group** | **Δ (MIC − Rest)** | **SE** | **95% CI** | **p** |
|---|---|---|---|---|---|---|
| Trapezius | Frequency (Hz) | CG | +0.192 | 0.135 | [-0.076, 0.459] | .158 |
|  | Frequency (Hz) | NPG | +0.197 | 0.135 | [-0.071, 0.465] | .147 |
|  | Stiffness (N/m) | CG | +5.214 | 3.487 | [-1.722, 12.151] | .139 |
|  | Stiffness (N/m) | NPG | +5.435 | 3.487 | [-1.502, 12.371] | .123 |
|  | Logarithmic Decrement | CG | +0.001 | 0.009 | [-0.016, 0.019] | .881 |
|  | Logarithmic Decrement | NPG | +0.023 | 0.009 | [0.006, 0.040] | **.010** |
|  | Relaxation time (ms) | CG | -0.132 | 0.231 | [-0.592, 0.328] | .569 |
|  | Relaxation time (ms) | NPG | -0.270 | 0.231 | [-0.729, 0.190] | .247 |
|  | Creep | CG | -0.012 | 0.012 | [-0.037, 0.012] | .322 |
|  | Creep | NPG | -0.014 | 0.012 | [-0.038, 0.010] | .261 |
| SCM | Frequency (Hz) | CG | +0.277 | 0.115 | [0.047, 0.506] | **.019** |
|  | Frequency (Hz) | NPG | +0.218 | 0.115 | [-0.012, 0.447] | .062 |
|  | Stiffness (N/m) | CG | +6.833 | 2.610 | [1.641, 12.026] | **.011** |
|  | Stiffness (N/m) | NPG | +3.012 | 2.610 | [-2.181, 8.204] | .252 |
|  | Logarithmic Decrement | CG | +0.007 | 0.019 | [-0.030, 0.044] | .716 |
|  | Logarithmic Decrement | NPG | -0.015 | 0.019 | [-0.052, 0.023] | .433 |
|  | Relaxation time (ms) | CG | -0.586 | 0.296 | [-1.175, 0.002] | .051 |
|  | Relaxation time (ms) | NPG | -0.475 | 0.296 | [-1.064, 0.114] | .112 |
|  | Creep | CG | -0.036 | 0.018 | [-0.071, -0.000] | **.047** |
|  | Creep | NPG | -0.021 | 0.018 | [-0.057, 0.014] | .231 |

**Note**.  Model-estimated contrasts (MIC − Rest) for healthy controls and participants with chronic neck pain. SCM = sternocleidomastoid; CG = healthy control group (n = 42); NPG = chronic neck pain group (n = 42); MIC = maximum intercuspation; Δ = model-estimated mean difference. Positive values indicate an increase from rest to MIC. Confidence intervals and p values are unadjusted. Contrasts were derived from separate linear mixed-effects models with participant-specific random intercepts. Bold p values indicate p < .05.

In the trapezius, the NPG showed a statistically significant increase in logarithmic decrement during MIC (estimated mean difference = 0.023, 95% CI [0.006, 0.040], p = .010). No statistically significant changes relative to rest were identified for frequency, stiffness, relaxation time, or creep in the NPG (all p > .05). In the CG, none of the five trapezius properties showed statistically significant changes between rest and MIC (all p > .05).

In the SCM, the CG showed statistically significant increases in frequency (estimated mean difference = 0.277 Hz, 95% CI [0.047, 0.506], p = .019) and stiffness (estimated mean difference = 6.833 N/m, 95% CI [1.641, 12.026], p = .011). A statistically significant reduction in creep was also identified (estimated mean difference = −0.036, 95% CI [−0.071, −0.000], p = .047). The reduction in relaxation time did not reach statistical significance (estimated mean difference = −0.586 ms, 95% CI [−1.175, 0.002], p = .051), and no statistically significant change was identified for logarithmic decrement (p = .716).

In the NPG, none of the five SCM properties showed statistically significant changes from rest to MIC (all p > .05).

These within-group contrasts describe changes associated with MIC within each group and should not be interpreted as evidence of differences in the magnitude of change between groups. All within-group contrasts are reported in Table 6, and the corresponding condition-specific measurements are illustrated in Figures 6 and 7.

#### 3.1.10. Sensitivity analyses of between-group changes

Visual inspection of residual-versus-fitted and quantile–quantile plots identified deviations from residual normality in selected MIC models, particularly in the tails of the residual distributions. Differences in residual dispersion were also observed across some model strata. The largest ratio between residual standard deviations was approximately 1.59, corresponding to trapezius relaxation time.

Sensitivity analyses of between-group differences in changes from rest to MIC were performed using participant-level stratified bootstrap resampling with 10,000 iterations and Yuen’s robust test based on 20% trimmed means. Holm’s step-down procedure was applied across the ten Yuen tests to account for multiple comparisons. Complete results are presented in Supplementary Table S6.

The bootstrap analyses yielded 95% confidence intervals that included zero for all ten outcomes. In the trapezius, the estimated between-group differences in mean change (NPG − CG) were 0.005 Hz for frequency (95% CI [−0.361, 0.370]), 0.220 N/m for stiffness (95% CI [−9.155, 9.518]), and 0.022 for logarithmic decrement (95% CI [−0.003, 0.045]). The corresponding differences were −0.137 ms for relaxation time (95% CI [−0.804, 0.469]) and −0.002 for creep (95% CI [−0.035, 0.032]).

In the SCM, the estimated between-group differences in mean change were −0.059 Hz for frequency (95% CI [−0.384, 0.255]), −3.821 N/m for stiffness (95% CI [−11.286, 3.310]), −0.022 for logarithmic decrement (95% CI [−0.072, 0.030]), 0.111 ms for relaxation time (95% CI [−0.732, 0.910]), and 0.014 for creep (95% CI [−0.034, 0.063]).

For trapezius logarithmic decrement, Yuen’s robust test indicated a nominal between-group difference in changes from rest to MIC (t(45.50) = 2.10, unadjusted p = .041). However, this difference was no longer statistically significant after Holm correction (adjusted p = .413). The corresponding group-by-condition interaction in the linear mixed-effects model was also not statistically significant (F(1, 82) = 3.12, p = .081), and the bootstrap confidence interval included zero.

No other outcomes showed statistically significant between-group differences in Yuen’s robust tests before adjustment (all unadjusted p > .05), and none of the ten comparisons remained statistically significant after Holm correction. The sensitivity analyses therefore provided no statistically significant evidence of between-group differences in changes from rest to MIC after adjustment for multiple comparisons. These analyses specifically addressed between-group differences in condition-related changes and should not be interpreted as direct assessments of the robustness of the significant main effects of condition.

#### 3.1.11. Exploratory between-group comparisons in a subsample

Exploratory between-group comparisons were conducted in a subsample of 36 participants, comprising 18 healthy controls (CG) and 18 participants with chronic neck pain (NPG), using Welch’s independent-samples t-tests.

The NPG exhibited significantly lower cervical extension than the CG (mean difference [CG − NPG] = 10.22°, 95% CI [1.90, 18.54], t(23.14) = 2.54, p = .018, Hedges’ g = 0.83). Significant between-group differences were also identified for right cervical rotation (mean difference = 8.17°, 95% CI [0.93, 15.40], t(34.00) = 2.29, p = .028, g = 0.75) and left cervical rotation (mean difference = 7.67°, 95% CI [1.41, 13.93], t(33.12) = 2.49, p = .018, g = 0.81). No statistically significant differences were identified for cervical flexion or right and left lateral flexion (all p > .05).

No statistically significant between-group differences were identified for tongue pressure at 50%, 80%, or 100% of MTP (all p > .05). Similarly, none of the myotonometric properties examined—muscle frequency, stiffness, logarithmic decrement, relaxation time, or creep—showed statistically significant between-group differences at rest or during the 50%, 80%, and 100% tongue-pressure conditions (all p > .05).

Complete descriptive statistics, mean differences, 95% confidence intervals, Welch’s t-test results, and Hedges’ g effect sizes are presented in Supplementary Table S7. All reported p values are unadjusted for multiple comparisons.

#### 3.1.12. Correlations between cervical range of motion, tongue pressure, and myotonometric properties

Spearman’s rank correlation analyses were performed separately in the CG (n = 18) and neck pain group (NPG; n = 18), using myotonometric measurements obtained during tongue-pressure tasks at 50% and 100% of MTP. A total of 308 correlations were examined, including 132 in the CG and 176 in the NPG. The correlation matrices, showing the original coefficients and unadjusted statistical significance, are presented in Figures 8 and 9. To assess the robustness of these exploratory associations to multiple testing, the Benjamini–Hochberg false discovery rate (FDR) procedure was applied separately within each group. Correlations remaining significant after adjustment are reported in Supplementary Table S8.

**Figure 8.**
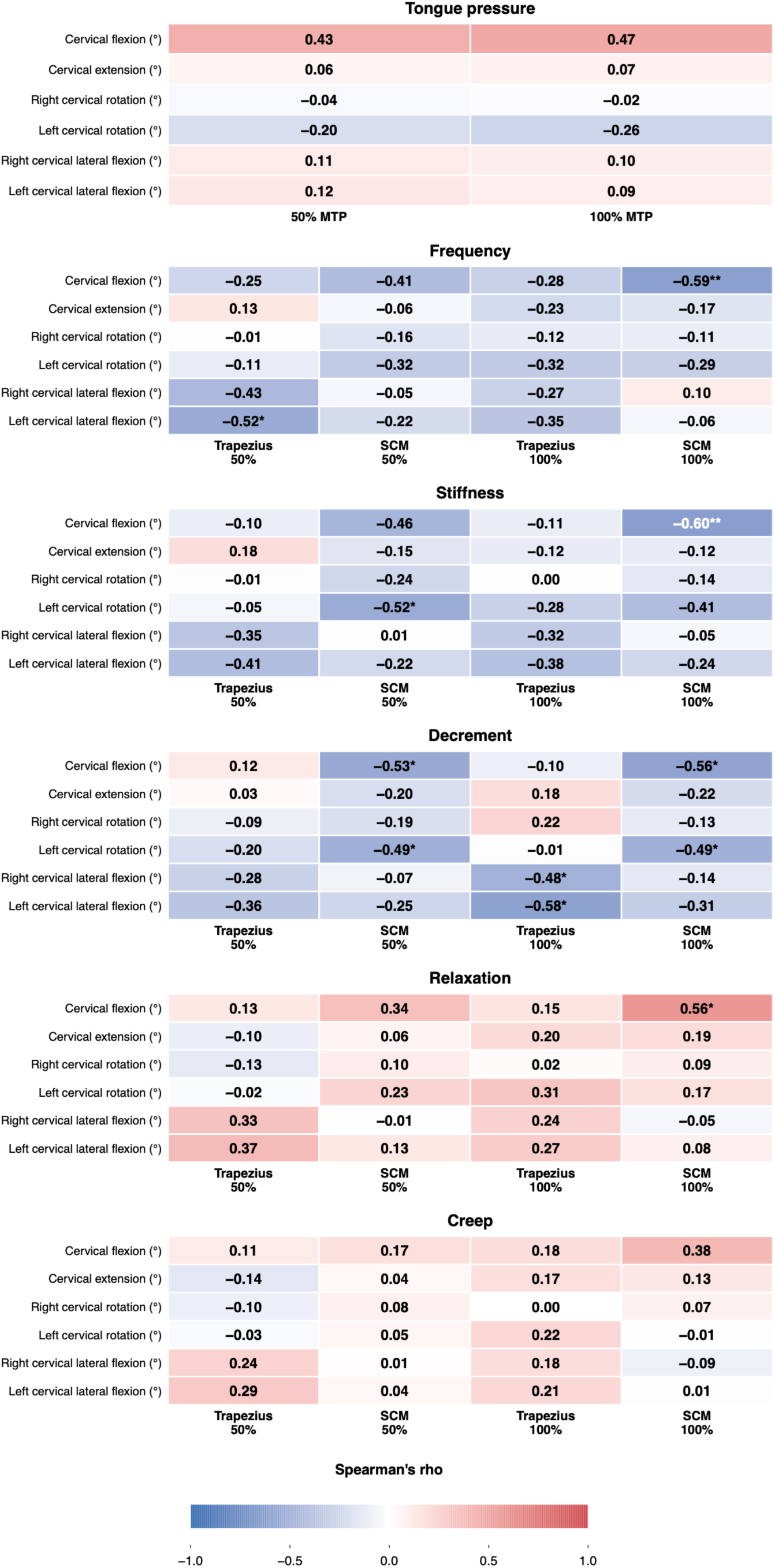
Spearman correlations in the control group. Heatmap showing Spearman’s rank correlation coefficients between cervical range of motion, tongue pressure, and myotonometric properties (frequency, stiffness, logarithmic decrement, relaxation time, and creep) during tongue-pressure tasks performed at 50% and 100% of maximum tongue pressure in the control group (n = 18). Blue represents negative correlations and red represents positive correlations, with color intensity reflecting the magnitude of Spearman’s ρ. Values within cells indicate correlation coefficients. Asterisks indicate nominal statistical significance based on unadjusted p-values: p < .05 (*), p < .01 (**), and p < .001 (***). None of the 132 correlations in the control group remained statistically significant after Benjamini–Hochberg false discovery rate adjustment. Results of the supplementary multiple-testing analysis are reported in Supplementary Table S8. SCM = sternocleidomastoid.

**Figure 9.**
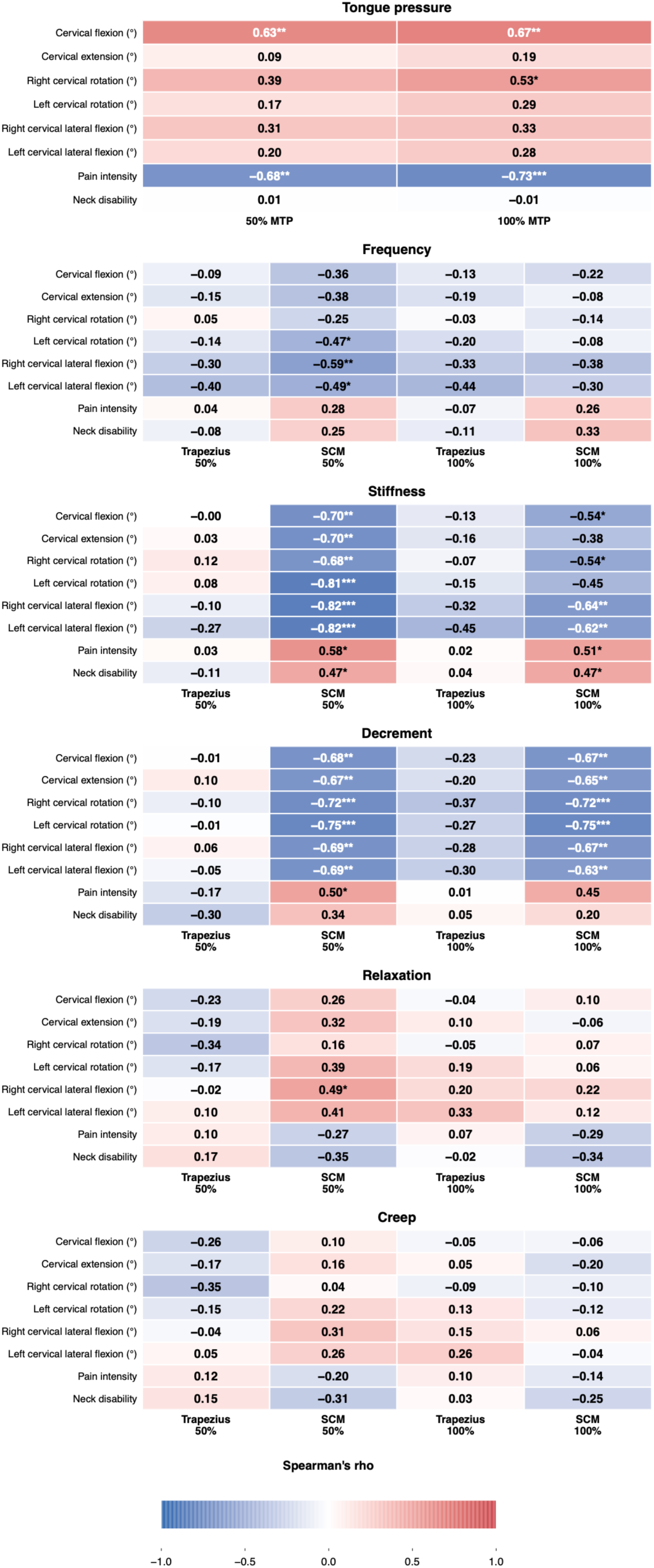
Spearman correlations in the neck pain group. Heatmap showing Spearman’s rank correlation coefficients between cervical range of motion, pain intensity, neck disability, tongue pressure, and myotonometric properties (frequency, stiffness, logarithmic decrement, relaxation time, and creep) during tongue-pressure tasks performed at 50% and 100% of maximum tongue pressure in the neck pain group (n = 18). Blue represents negative correlations and red represents positive correlations, with color intensity reflecting the magnitude of Spearman’s ρ. Values within cells indicate correlation coefficients. Asterisks indicate nominal statistical significance based on unadjusted p-values: p < .05 (*), p < .01 (**), and p < .001 (***). Following Benjamini–Hochberg false discovery rate adjustment, 24 of the 176 correlations remained statistically significant. These associations are reported in Supplementary Table S8. SCM = sternocleidomastoid.

In the NPG, cervical range of motion was negatively associated with SCM stiffness and logarithmic decrement across several movement directions. Particularly strong associations were identified between SCM stiffness at 50% MTP and left cervical rotation (ρ = −0.81), right cervical lateral flexion (ρ = −0.82), and left cervical lateral flexion (ρ = −0.82). All three associations remained statistically significant after FDR adjustment (adjusted p = .0024 for each comparison). These findings indicate that greater cervical mobility was associated with lower SCM stiffness under this experimental condition.

Negative associations were also observed between cervical mobility and SCM logarithmic decrement at both tongue-pressure intensities. Left cervical rotation was correlated with logarithmic decrement at 50% and 100% MTP (ρ = −0.75 for both conditions; FDR-adjusted p = .0109). Additional correlations involving cervical flexion, extension, rotation, and lateral flexion remained statistically significant after adjustment, indicating that these associations were not restricted to a single cervical movement direction.

Pain intensity was negatively associated with tongue pressure at both experimental intensities. The correlation coefficients were ρ = −0.68 at 50% MTP (FDR-adjusted p = .0238) and ρ = −0.73 at 100% MTP (FDR-adjusted p = .0152). Higher pain intensity was therefore associated with lower tongue-pressure values in this subsample. Positive correlations were also observed between neck disability and SCM stiffness at both intensities (ρ = 0.47); however, these associations did not remain statistically significant after FDR adjustment.

In the CG, cervical flexion was negatively correlated with SCM frequency (ρ = −0.59, unadjusted p < .01) and stiffness (ρ = −0.60, unadjusted p < .01) at 100% MTP. Additional nominally significant negative correlations were identified between cervical mobility and logarithmic decrement, particularly for cervical flexion and rotation. Nevertheless, none of the 132 correlations examined in the CG retained statistical significance after FDR adjustment. These results indicate that the associations initially identified in this group were not sufficiently robust to remain statistically significant under the applied multiple-testing procedure.

Before adjustment, 11 correlations in the CG and 36 in the NPG met the nominal significance threshold of p < .05. Following the Benjamini–Hochberg procedure, 24 correlations remained significant in the NPG, whereas none survived in the CG. The retained associations predominantly involved cervical range of motion and SCM mechanical properties, particularly stiffness and logarithmic decrement, together with associations between pain intensity and tongue pressure (Supplementary Table S8). Although several correlations were relatively strong, these findings should be interpreted cautiously because of the exploratory nature of the analyses, the limited sample size within each group, and the interdependence of the measurements. The observed associations do not establish causal relationships and require confirmation in independent samples.

### 3.2. DISCUSSION

The principal finding of this study was that tongue-pressure tasks and MIC were associated with changes in cervical muscle mechanical properties in both individuals with chronic neck pain and healthy controls. During tongue-pressure tasks, condition-related responses were observed across most muscle-property combinations, particularly for frequency, stiffness, relaxation time, and creep. MIC was associated with a more restricted pattern of changes, while evidence of differences in task-related mechanical responses between groups was limited. These observations indicate that cervical tissues respond mechanically during orofacial motor activity, although the contribution of active muscle recruitment and other physiological mechanisms cannot be established from myotonometric measurements alone. The exploratory findings further suggest relationships between cervical mobility, selected mechanical properties, and tongue-pressure performance, particularly among participants with chronic neck pain.

The mechanical changes observed during tongue-pressure tasks are consistent with previous investigations describing functional relationships between the cervical and hyolingual systems. Paris-Alemany et al. (2021) demonstrated that craniocervical posture influenced tongue strength and endurance, suggesting that lingual motor performance is sensitive to cervical positioning. Pérez-González et al. (2025) subsequently examined the effects of passive cervical immobilization on tongue mobility and strength in 33 healthy adults. Although cervical immobilization did not produce a significant overall change in tongue strength, the authors identified differences associated with tongue region and changes in anterior tongue mobility. Their subgroup analyses also suggested that the middle region of the tongue may be particularly sensitive to cervical restriction. These observations indicate that the relationship between cervical configuration and tongue performance is dependent on the specific function and anatomical region examined. The present study extends this line of investigation by examining cervical tissue mechanical behavior during lingual force production rather than assessing how cervical restriction modifies tongue performance.

Experimental evidence obtained using elastography provides a more direct comparison with our mechanical findings. Sherwin et al. (2024) investigated the effects of tongue-to-palate pressure on deep neck flexor stiffness in healthy adults. Tongue pressure increased deep cervical muscle stiffness under specific resting conditions, whereas the response was less evident during cervical tasks that already required muscular engagement. This suggests that the mechanical response associated with lingual activity may depend on the pre-existing demands imposed on the cervical musculature. Our results are compatible with this interpretation, as tongue-pressure conditions were associated with increases in frequency and stiffness and reductions in relaxation time and creep. Nevertheless, Sherwin et al. examined deep cervical flexors using shear wave elastography, whereas the present study evaluated superficial SCM and trapezius muscles using myotonometry. Differences in muscle anatomy and measurement principles limit direct comparisons of the magnitude and physiological meaning of the observed responses.

Additional evidence comes from Pérez-González et al. (2026), who investigated electromyographic responses in cervical, masticatory, and hyoid muscles during tongue-palate pressure tasks performed with and without craniocervical motion restriction. Cervical restriction was associated with increased activity in several hyoid and masticatory muscles, while the responses of the cervical muscles were less consistent. In particular, trapezius electromyographic activity did not show a significant increase under the experimental conditions examined. This finding is relevant because it indicates that functional interaction between cervical and orofacial motor systems does not necessarily involve a generalized increase in electrical activity across all cervical muscles. The differences between that investigation and the present results may be related to the experimental manipulation, the muscles examined, and the measurement techniques employed. Electromyography characterizes electrical muscle activity, whereas myotonometry assesses the response of superficial tissues to a mechanical impulse. Changes in myotonometric stiffness or frequency therefore cannot be assumed to represent proportional changes in motor-unit recruitment.

The broader literature on mandibulocervical coordination provides a physiological context for these observations. Eriksson et al. (1998) documented concomitant mandibular and head-neck movements during jaw opening and closing, while Eriksson et al. (2000) described coordinated movement patterns during rhythmic jaw activities. Häggman-Henrikson and Eriksson (2004) further demonstrated that head movements during chewing varied according to the size and texture of the food bolus. These studies suggest that cervical participation in orofacial behavior is influenced by the demands of the motor task. Although they primarily examined movement coordination rather than tissue mechanical properties, they support the interpretation that cervical muscle behavior may change during oral motor activity even when the cervical musculature is not the primary agonist of the task.

The direction of the mechanical responses observed in the present study requires careful interpretation. During tongue-pressure tasks, muscle frequency and stiffness generally increased, whereas relaxation time and creep decreased. These changes are compatible with a modification of tissue mechanical behavior during sustained force production. However, the five myotonometric parameters describe different properties and cannot be considered interchangeable indicators of muscle activation. Frequency characterizes the natural oscillation of the tissue following mechanical stimulation, stiffness reflects resistance to deformation, logarithmic decrement describes oscillation damping, and relaxation time and creep characterize aspects of viscoelastic behavior. Their physiological interpretation depends on the mechanical conditions under which measurements are obtained. Wolff et al. (2022), who investigated muscle activity and elasticity during functional reaching in individuals with and without chronic neck pain, demonstrated the importance of examining mechanical and electromyographic responses as related but distinct outcomes. Without simultaneous electromyographic recordings, the present study cannot determine the contribution of active cervical muscle recruitment to the observed changes.

The interpretation of these mechanical responses also depends on the measurement properties of myotonometry. Previous investigations have documented differences in reliability and measurement error across cervical muscles and myotonometric parameters (Taş et al., 2021; Lettner et al., 2024; Koterba & Saulicz, 2025). This variability is relevant because statistically significant changes may not necessarily exceed the measurement error associated with a particular parameter. Although standardized procedures and repeated measurements were used, the present analyses did not establish whether the observed responses exceeded parameter-specific measurement error under the experimental conditions examined. The physiological magnitude and potential clinical relevance of these changes therefore remain uncertain.

The effects of tongue-pressure conditions were not accompanied by consistent differences between individuals with chronic neck pain and healthy controls. No significant group-by-condition interactions were identified for frequency, stiffness, relaxation time, or creep. Although several within-group comparisons reached statistical significance in the neck pain group, the presence of a significant response in one group and a nonsignificant response in another does not establish a between-group difference. The mixed-effects models therefore provide limited evidence that chronic neck pain modified the mechanical response to lingual force production. This finding is particularly relevant because it suggests that the mechanical changes associated with tongue-pressure tasks may reflect functional responses that are also present in asymptomatic individuals.

The absence of consistent group-specific responses should be interpreted within contemporary models of motor adaptation to pain. Hodges and Tucker (2011) proposed that pain-related adaptations involve changes in the distribution of muscle activity and movement strategies, with considerable variability between individuals and tasks. Their framework challenges the assumption that pain produces a uniform increase in muscle activation or stiffness. Falla and Farina (2008) similarly described alterations in cervical neuromuscular control associated with experimental and clinical pain, while Boudreau and Falla (2014) demonstrated changes in muscle activation patterns during sudden movements in individuals with chronic neck pain. These findings indicate that pain-related adaptations may become apparent under particular motor demands without necessarily producing consistent differences in isolated mechanical measurements. In the present study, the cervical muscles were evaluated during tasks primarily involving lingual force production. It remains possible that differences in motor coordination, recruitment strategies, or movement variability were present but were not captured by the selected myotonometric outcomes. This explanation remains hypothetical because the study did not directly assess these aspects of motor control.

The findings also require comparison with previous studies of cervical muscle stiffness in symptomatic populations. Opara and Kozinc (2023), in a systematic review and meta-analysis, reported greater upper trapezius stiffness in individuals with chronic neck pain than in asymptomatic controls, although the pooled effect was relatively small and the available evidence was heterogeneous. The absence of significant main group effects in our study differs from this pooled finding. Several methodological considerations may contribute to the discrepancy. Mechanical properties vary with posture, anatomical measurement site, contraction state, and the technique used to characterize tissue behavior. Differences in symptom duration, pain intensity, disability, and participant characteristics may also influence between-group estimates. Importantly, an elevated resting stiffness and an altered response to an active task represent different phenomena. The present results did not identify consistent group differences across the experimental conditions, but nonsignificant comparisons should not be interpreted as demonstrating equivalence between symptomatic and asymptomatic participants.

Logarithmic decrement exhibited a less consistent response than the other myotonometric parameters. Significant condition effects were identified in the right SCM and both trapezius muscles, whereas the left SCM did not show a significant condition effect. A group-by-condition interaction was observed for the right trapezius, although the corresponding comparisons of changes from rest did not establish a consistent group-specific pattern. A nominal condition-specific difference was also identified in the left trapezius at 80% MTP despite the absence of a significant interaction for that muscle. The relatively small interaction effect and the number of comparisons performed limit the interpretation of these isolated findings. Logarithmic decrement reflects the damping of mechanically induced tissue oscillations, and its behavior may differ from parameters such as frequency and stiffness. The reliability findings reported by Taş et al. (2021), Lettner et al. (2024), and Koterba and Saulicz (2025) provide an additional reason to interpret isolated changes in viscoelastic parameters cautiously. The present data do not allow the contributions of physiological variability and measurement characteristics to be separated.

The second objective concerned cervical muscle mechanical responses during maximum intercuspation. Significant condition effects were identified for frequency and stiffness in both the trapezius and SCM, with additional reductions in relaxation time and creep in the SCM. These results are consistent with previous evidence of cervical muscle involvement during mandibular force production. Ehrlich et al. (1999) demonstrated increased electromyographic activity in cervical and trunk muscles during jaw clenching, providing experimental evidence that forceful mandibular contraction may be accompanied by recruitment of muscles outside the primary masticatory system. Giannakopoulos et al. (2013) subsequently examined neuromuscular interactions between jaw and cervical muscles during controlled clenching and showed that cervical coactivation varied according to the magnitude and direction of the applied bite force. These investigations support the interpretation that MIC may impose mechanical and neuromuscular demands on the cervical region.

The present results extend this evidence by demonstrating changes in cervical myotonometric properties during maximum intercuspation. The increases in frequency and stiffness observed in both muscles are compatible with changes in tissue mechanical behavior during mandibular force production. However, the electromyographic findings reported by Ehrlich et al. (1999) and Giannakopoulos et al. (2013) cannot be used to infer the magnitude of cervical activation in the present study. The relationship between electrical activity and mechanical stiffness is influenced by several factors, including tissue loading and muscle geometry. The absence of direct electromyographic recordings therefore limits conclusions concerning the neuromuscular mechanisms responsible for the observed responses.

The pattern of changes during MIC differed descriptively from that observed during tongue-pressure tasks. Frequency and stiffness were affected in both cervical muscles, whereas significant changes in relaxation time and creep were restricted to the SCM. During tongue-pressure tasks, condition effects were more consistently observed across the measured parameters. These differences may reflect the distinct mechanical requirements of lingual pressure production and mandibular clenching. Nevertheless, the two tasks were not directly compared in a model designed to estimate differences between task types. The available findings therefore do not establish that tongue-pressure tasks produce greater cervical mechanical responses than maximum intercuspation. Differences in contraction duration, force direction, relative effort, and stabilization demands remain possible explanations that require direct experimental evaluation.

No significant group-by-condition interactions were identified for the maximum-intercuspation outcomes. Although the neck pain group exhibited a significant within-group increase in trapezius logarithmic decrement, this finding was not accompanied by a statistically significant interaction. The sensitivity analyses supported a cautious interpretation. A nominal between-group difference in trapezius decrement change was identified using Yuen’s robust test, but it did not remain significant after Holm adjustment. The bootstrap confidence intervals for the between-group differences in change included zero across all ten outcomes. These findings provide no statistically significant evidence that chronic neck pain systematically modified the mechanical response to maximum intercuspation. However, the absence of significant differences does not establish equivalence, particularly because the study was not designed to test predefined equivalence margins.

The third objective examined associations between cervical function, tongue pressure, myotonometric properties, and clinical characteristics. In the exploratory subsample, individuals with chronic neck pain demonstrated reduced cervical extension and bilateral rotation compared with healthy controls. These differences were accompanied by negative associations between cervical range of motion and SCM stiffness during tongue-pressure tasks. The strongest correlations were observed at 50% MTP, particularly for cervical lateral flexion. Negative correlations were also identified between cervical mobility and SCM logarithmic decrement. These findings suggest that cervical mechanical properties measured during an orofacial task may be related to cervical movement characteristics in symptomatic individuals. Their cross-sectional nature prevents conclusions about the direction of the relationship. Increased stiffness could be associated with restricted mobility, reflect an adaptation to movement limitation, or be related to other characteristics of the participants.

The clinical relevance of these associations can be considered in relation to evidence linking cervical pain and orofacial dysfunction. Bizzarri et al. (2026), in a systematic review and meta-analysis, reported a higher prevalence of temporomandibular disorders among individuals with neck pain and/or cervicogenic headache, together with differences in selected orofacial outcomes. The review identified an odds ratio of 3.64 for the association between neck pain and temporomandibular disorders, although the certainty of evidence was very low. These findings provide epidemiological support for an association between cervical and orofacial conditions, but they do not establish that cervical pain causes changes in tongue strength or cervical muscle mechanical properties. The present study investigated a different population and a different set of outcomes. Nevertheless, the clinical associations observed in the exploratory analyses are compatible with the possibility that cervical symptoms and orofacial motor performance share functional determinants.

Pain intensity was negatively correlated with tongue pressure at 50% and 100% MTP in the neck pain group. This finding warrants particular attention because the main sample also demonstrated lower absolute tongue-pressure values in the neck pain group at the 80% and 100% conditions. Arvanitidis et al. (2025), in a systematic review and meta-analysis, examined the influence of pain on force control and reported evidence of impaired force steadiness in clinical pain populations. Although force steadiness differs from tongue-pressure magnitude, the review supports the broader proposition that pain may be associated with changes in motor performance. The experimental findings of Pérez-González et al. (2025) further indicate that cervical restriction can influence selected aspects of tongue mobility and region-specific strength, while Pérez-González et al. (2026) demonstrated changes in orofacial muscle recruitment under cervical restriction conditions. These investigations provide plausible physiological contexts for the observed associations, but the present data cannot determine whether pain directly influenced tongue-force production. Differences in effort, cervical stabilization, or other unmeasured factors may have contributed to the results.

Positive correlations were observed between neck disability and SCM stiffness during tongue-pressure tasks in the neck pain group. However, these associations did not remain statistically significant after Benjamini–Hochberg FDR adjustment and should therefore be interpreted as nominal exploratory findings. In contrast, several associations involving cervical mobility and SCM mechanical properties retained statistical significance after correction. Although nominally significant correlations were also identified in healthy controls, none survived FDR adjustment in that group. This difference in the number of retained associations does not establish that the underlying correlations differ significantly between groups, as no direct between-group comparisons of correlation coefficients were performed. The exploratory findings do not establish the clinical utility of myotonometric measurements as markers of pain severity or neck disability, and their reproducibility requires evaluation in independent samples.

Several methodological considerations influence the interpretation of the findings. The study included equal numbers of individuals with chronic neck pain and healthy controls and used repeated measurements across standardized experimental conditions. The assessment of five mechanical properties in bilateral cervical muscles allowed the response to orofacial tasks to be characterized beyond stiffness alone. Mixed-effects models accounted for repeated observations within participants, and the maximum-intercuspation analyses were supplemented by bootstrap and robust sensitivity procedures. These features strengthen the characterization of condition-related responses. Nevertheless, the mechanical measurements were obtained from superficial tissues and cannot isolate the contributions of individual anatomical structures or distinguish passive tissue behavior from active neuromuscular recruitment. The findings of Sherwin et al. (2024) and Pérez-González et al. (2026), obtained using elastography and electromyography, respectively, illustrate the complementary information that could be gained from simultaneous measurement techniques.

The reliability literature also identifies an important limitation. Taş et al. (2021), Lettner et al. (2024), and Koterba and Saulicz (2025) reported variability in the reliability and measurement error of myotonometric parameters across muscles and experimental conditions. Although the present study identified numerous statistically significant condition effects, the magnitude of these changes was not systematically evaluated against parameter-specific minimum detectable changes established under comparable active-task conditions. This restricts conclusions about whether all statistically significant differences represent changes that are sufficiently large to exceed measurement variability. Future experimental protocols should incorporate direct estimates of measurement error under the same contraction conditions used for the primary analyses.

The groups also differed in sex distribution, with a higher proportion of women in the neck pain group. Sex was not included as a covariate in the final mixed-effects models, and residual confounding cannot therefore be excluded. The study involved multiple myotonometric outcomes and numerous statistical comparisons. Although multiplicity adjustments were applied to specified post hoc analyses, the omnibus tests across separate models were not jointly adjusted. The exploratory analyses were particularly vulnerable to multiplicity because of the large number of correlations examined in a relatively small sample. The results should therefore be interpreted according to the consistency of the observed patterns rather than the statistical significance of isolated comparisons.

Future investigations should examine cervical mechanical responses alongside electromyographic activity and direct measures of motor coordination to clarify the mechanisms underlying the observed changes during orofacial tasks. Longitudinal and experimental designs would also help determine whether the associations between cervical mobility, tongue-pressure performance, and myotonometric properties reflect stable functional relationships or vary according to pain-related adaptations. Particular attention should be given to measurement error under active contraction conditions and to the potential influence of individual characteristics on cervical muscle responses.

## 4. CONCLUSIONS

Tongue-pressure tasks and MIC were associated with changes in the mechanical properties of the SCM and trapezius muscles in individuals with and without chronic neck pain. These responses were more consistently observed during tongue-pressure tasks, although the experimental conditions did not provide consistent evidence of different cervical mechanical responses between groups. Exploratory associations involving cervical mobility, tongue pressure, and selected myotonometric properties suggest relationships between cervical and orofacial function that warrant further investigation. The findings indicate that cervical mechanical responses accompany orofacial motor activity, but their underlying mechanisms and clinical relevance remain to be established.

## 5. Data availability

The datasets generated and analyzed during the current study are available from the corresponding author upon reasonable request.

## 6. Conflicts of interest

The authors declare no conflicts of interest.

## 7. Funding statement

This research received no specific grant from any funding agency in the public, commercial, or not-for-profit sectors.

## Notes

### Competing Interest Statement

The authors have declared no competing interest.

### Author Declarations

Ethics committee/IRB of Centro Superior de Estudios Universitarios La Salle gave ethical approval for this work (CSEULS-PI 010/2020).

